# A disaggregated dynamic model of pulmonary and extrapulmonary tuberculosis for vaccine and booster-schedule evaluation: VPM1002 in India as a case study

**DOI:** 10.64898/2026.07.31.26359434

**Authors:** M Revathy, Vidya Niranjan, Akshay Uttarkar, Varun Swaminathan, Alessandro Carrese, Aditi Bhargava

## Abstract

**Introduction:** Extrapulmonary tuberculosis (EPTB) is rising in India and carries higher case fatality than pulmonary tuberculosis (PTB). A Phase III trial of VPM1002 missed its primary composite endpoint, but secondary endpoints showed a stronger EPTB signal (vaccine efficacy 42·3%, 95% CI −9·1 to 69·4). We identified no published model resolving tuberculosis by disease type, nutritional and socioeconomic status, and dose event against a dynamic baseline.

**Methods:** We built a deterministic model with 1,152 population compartments (72 strata by age, HIV status, body-mass index and socioeconomic status), calibrated to WHO India incidence and mortality for 2015–2024. Separate PTB and EPTB effectiveness posteriors, derived by Bayesian evidence synthesis of the trial’s per-protocol and modified intention-to-treat estimates, were propagated through 1,000 Monte Carlo iterations. Three doses were modeled at 10-year intervals assuming 3 or 10 years of protection. Economic analysis used societal and health-system perspectives with 3% discounting.

**Results:** Posterior median effectiveness was higher against EPTB (42·9%, 95% credible interval [CrI] −5·6 to 69·1) than PTB (14·0%, −19·0 to 37·9). Per 1,000,000 targeted under 10-year protection, three doses averted 5,218 cases (95% uncertainty interval 153–10,157), 682 deaths (24–1,255) and 9,463 disability-adjusted life-years (DALYs; 328–17,404); under 3-year protection, 1,961 cases and 3,708 DALYs. EPTB accounted for about two-thirds of averted deaths and DALYs but about half of averted cases. Under 10-year protection, the first booster added 46% more cases and 34% more DALYs at an incremental benefit–cost ratio of 19·1 (initial dose 20·7); the second booster added 8% more cases and 4·5% more DALYs at 6·8. Median gross incremental cost-effectiveness ratios were US$329 (10-year) and US$832 (3-year) per DALY averted.

**Conclusion:** Composite-endpoint models cannot show this dissociation. A booster a decade later was nearly as economically efficient but averted less disease. This is an adolescent and adult booster program, not a replacement for infant BCG.

**What is already known on this topic:**

- TB vaccine modeling studies for India and other high burden settings have relied on aggregate single efficacy estimates that combine pulmonary and extrapulmonary disease into a single endpoint.
- Published models have evaluated vaccines against static programmatic baselines and a single course intervention, so the marginal value of revaccination has rarely been quantified.

**What this study adds:**

- This 1,152 population compartment model tracks pulmonary and extrapulmonary TB as separate compartments with independently calibrated natural history and separately propagated Bayesian effectiveness posteriors.
- Vaccination history is preserved through disease treatment and recovery, so people infected after vaccination remain correctly identified as previously vaccinated and eligible for later doses; this becomes relevant only once boosting is modeled
- Extrapulmonary disease accounts for approximately 2/3 of averted deaths and DALYs against roughly half of averted cases, a dissociation that a composite endpoint model cannot provide.
- Seed-matched paired Monte Carlo makes each dose event’s marginal contribution directly estimable rather than inferential.

**How this study might affect research, practice or policy:**

- Policy assessments anchored to composite endpoints, or assuming a single lifetime dose, may underestimate this vaccine’s population value.
- A single booster a decade after the initial dose is close to as economically efficient as the initial dose; a second booster is materially less so within a horizon to 2050.
- The framework is country- and candidate-agnostic and can be recalibrated for other high-burden settings and other vaccine candidates, including those evaluated in other Phase III trials.
- The schedule evaluated here is an adolescent and adult booster program and does not bear on infant BCG policy.

## Introduction

Tuberculosis remains among the leading infectious causes of death globally, with 1·2 million deaths annually and India contributing approximately 25% of global incidence.^1 2^ The WHO End TB Strategy has driven meaningful reductions in PTB, but one dimension of the epidemic is moving the other way. EPTB incidence has been rising in India, and in a ten-year European cohort it held steady while pulmonary incidence fell, so its share of the total rose; it also remains substantially underdiagnosed.^3–5^ Microbiological confirmation is achieved in only approximately 16% of EPTB cases, because bacillary load in extrapulmonary tissues is low,^6^ culture methods are poorly standardized for non-respiratory specimens, and diagnostic access is limited. The commonest manifestations, namely, lymphatic, pleural, meningeal and osteoarticular TB carry higher hospitalization costs, greater disability and higher case fatality than pulmonary disease, and EPTB is disproportionately prevalent in HIV-positive, immunocompromised and malnourished people, the groups least well served by detection-based programs. While tuberculosis preventative treatment (TPT) provided sustained protection in medium-to-low-burden settings, the protection waned rapidly within a year in high burden settings such as India, attributed largely to reinfection.^7^ Vaccines for adolescents and adults are widely recognized as critical to future TB control, and several candidates have been evaluated recently, including M72/AS01ᴇ, BCG revaccination, MVA85A and VPM1002.^8–12^

The PreVenTB phase III trial enrolled 12,717 healthy household contacts aged ≥6 years across eighteen sites in India.^13^ Here, we used only the VPM1002-versus-placebo comparison (modified intention-to-treat 4,207 against 4,223; per-protocol 3,860 against 3,845). The primary composite endpoint, microbiologically confirmed all-TB, gave a vaccine efficacy of 16·9% (−13·3 to 39·1), the confidence interval crossing zero. Disaggregated, the EPTB signal was substantially stronger: 42·3% (−9·1 to 69·4) in the mITT population and 50·4% (0·8 to 75·2, p=0·04) per protocol.^13^ This asymmetry is biologically coherent. The listeriolysin O modification in recombinant VPM1002 BCG vaccine enables phagosomal escape and MHC class I antigen presentation.^14^ CD8 T-cell and trained-innate responses are the proposed downstream mechanism; such responses would not depend on prior *Mycobacterium tuberculosis* sensitization, and EPTB predominantly affects immunocompromised hosts, who are frequently IGRA-negative. That link is a hypothesis consistent with the endpoint asymmetry rather than something this analysis tests.

Several transmission models have been used to translate TB vaccine trial efficacy into population-level policy guidance. We identified two structural gaps in the current TB vaccine modelling literature. First, no published model tracks PTB and EPTB as separate disease compartments. Every cost-effectiveness estimate for TB vaccines in India and other high-burden settings,^15–22^ has been built on composite endpoints that treat EPTB as background noise within a PTB-centric framework. For vaccines with predominant PTB and negligible EPTB efficacy, that approach is reasonable; for VPM1002, whose largest point estimate of efficacy is EPTB-specific, it is not. The current models conflate two disease types with different case fatality, disability weights and demographic distributions, obscuring where benefit falls. Second, vaccine impact is almost always modelled as a continuous rollout or a single lifetime dose, so the marginal value of revaccination is rarely ascertained, even though any deployment of a waning vaccine must guide whether and when to boost.

Here, we developed a dynamic transmission model of PTB and EPTB stratified by nutritional and socioeconomic status. Disaggregated PTB and EPTB were modelled as separate compartments with independently calibrated natural history, separately propagated effectiveness posteriors, separately reported health outcomes and economics, and an explicit multi-dose schedule whose marginal contribution can be estimated rather than inferred. We used India as the case study because the trial was conducted there, so the effectiveness estimates come from the population being modeled and need no assumption about transfer between countries, and because EPTB incidents in India is rising. Additionally, we constructed a dynamic National Tuberculosis Elimination Program (NTEP) baseline calibrated to observed 2015–2024 data, following the Stop TB Partnership sequential intervention framework,^19^ so vaccine impact is a marginal increment over a realistically declining trajectory rather than a frozen 2015 benchmark. Dual-perspective outputs are reported in a format usable by India’s National Technical Advisory Group on Immunization and Health Technology Assessment in India and directly comparable to published ICER-based analyses,^16–18 21^ enabling a like-for-like evaluation of what is gained when EPTB is tracked separately rather than aggregated. The program modelled here vaccinates people aged 6 years and older. It is a booster strategy for adolescents and adults and not a replacement for neonatal BCG; the model contains no age stratum below 6 years. Although calibrated to India, the demographic stratification, dynamic baseline and evidence-synthesis pipeline are agnostic to geography and vaccine candidate: recalibration needs country-specific epidemiological and program inputs, and a different vaccine needs only substitution of its own trial-derived Bayesian posteriors, such as M72/AS01ᴇ and other future TB vaccines.

## Methods

### Model structure

We developed a deterministic compartmental model of TB disaggregated by clinical form into PTB, EPTB and concurrent PTB–EPTB disease (MIX), which differ in transmissibility, case fatality and disability burden. All three contributed to the force of infection, at disease-specific infectiousness weights of 1·00, 0·30 and 0·65. Separate vaccinated-latent compartments allowed vaccine effects on reactivation to be modeled independently of effects on primary infection. The population was stratified into age band a ∈ {1,2,3,4} (6–14, 15–30, 31–55, 56–80 years), HIV status h ∈ {negative, positive}, body-mass-index category b ∈ {low, normal, high}, and socioeconomic status s ∈ {low, middle, high}, giving 4 × 2 × 3 × 3 = 72 strata. Within each stratum, 30 disease states of three types were tracked. Sixteen were population compartments, each held people at any point in time. Seven counted events: incident cases and deaths by disease type, and doses administered. The remaining seven recorded what proportion of the people held in the disease, treatment and recovery compartments had been vaccinated before infection; because these duplicate a subset of their parent compartments, they were excluded from population totals to prevent double counting (figure 1, online supplemental methods and table S1). The system was integrated as a set of ordinary differential equations using LSODA, over a burn-in period followed by the 2015–2050 reporting window (online supplemental table S2).

**Figure 1.**
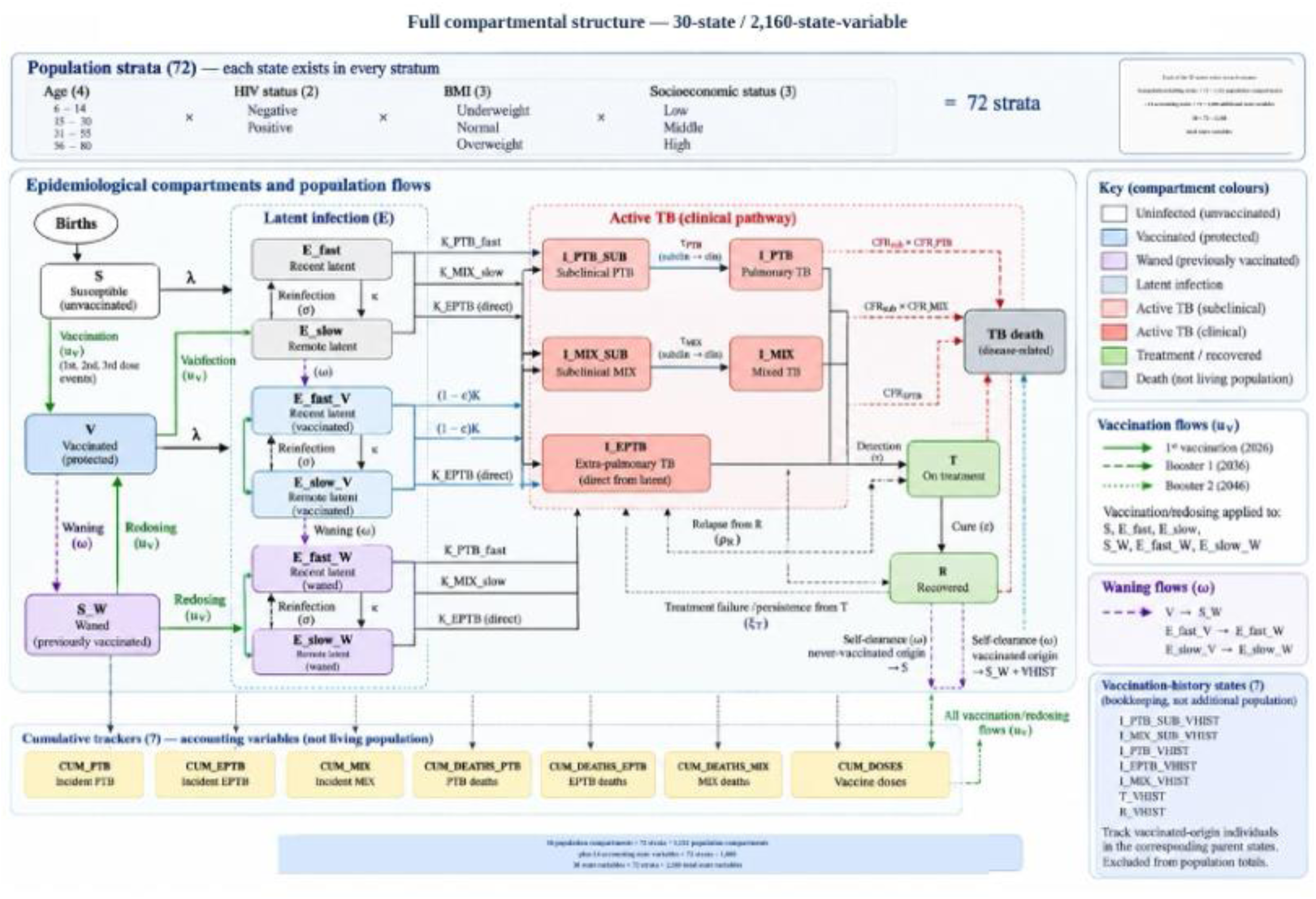
A deterministic dynamic 1,156 population compartmental model of PTB and EPTB stratified by nutritional and socioeconomic status for vaccine and booster-schedule evaluation. Strata are defined by age (4 bands), HIV status (2 states), BMI (3 states), and socioeconomic status (3 states). Of the 30 states, 16 carry real population. Seven cumulative trackers count events rather than individuals; incident PTB, EPTB, and MIX cases, deaths from each disease type, and total doses delivered. Seven vaccination-history states record how many of the people already counted in the subclinical, clinical, treatment, and recovered compartments were vaccinated before infection; these duplicate a subset of their parent compartments and are excluded from population totals to avoid double counting. Susceptible and latent pools are each split three ways by vaccination status: never vaccinated, vaccinated with protection intact, and previously vaccinated with protection waned. Progression from the vaccinated latent pools carries the blocking multiplier (1 − ε_prog), applied separately to pulmonary and extrapulmonary disease. Waned pools progress at the unprotected rate but remain eligible for the next scheduled dose, which is how the 2036 and 2046 boosters are applied. Self-clearance from the recovered pool is routed by origin: people of vaccinated origin return to the waned susceptible compartment rather than the never vaccinated one, so the vaccination history survives a full disease episode. Pulmonary and mixed disease pass through a subclinical stage before clinical presentation; extrapulmonary disease is modeled without one. PTB = pulmonary tuberculosis; EPTB = extrapulmonary tuberculosis; MIX = concurrent pulmonary and extrapulmonary diseases.

Two structural features distinguished this model from other TB vaccine models. First, a subclinical stage that lay between latent infection and clinical pulmonary or concurrent disease, with its own progression rate, self-cure rate, case fatality and infectiousness was incorporated into the model. This is because between 36% and 80% of bacteriologically confirmed TB in population prevalence surveys is subclinical;^23^ incidence was counted at entry to that stage. Extrapulmonary disease has no subclinical stage, because it is not detected by the symptom- and sputum-based screening that defines subclinical pulmonary TB operationally. Second, vaccination history was preserved through a full disease episode: waning of natural immunity in the vaccinated-origin recovered pool returned people to the susceptible compartment for those whose protection had lapsed, where they remained eligible for the next scheduled dose, rather than to the never-vaccinated susceptible pool. This is irrelevant in a single-dose analysis but relevant once boosting is modeled. Claude Code (Anthropic [Opus 5.5]; Aseesa Inc.’s institutional license) was used to assist with coding. All code and model outputs were independently checked and verified by the authors (RM, VN, VM, AU, and AC).

### Calibration and baseline projection

Calibration was designed in two layers. Layer 1 fitted four-parameter logistic functions to India’s observed case-detection rate (CDR) for 2015–2024 and treatment-success rate (TSR) for 2015–2023, the latest year available, with the calibration origin at 2015 (online supplemental figure S1).^24 25^ Layer 2 estimated 14 free natural-history and program parameters by differential evolution; search bounds and calibrated values are given in online supplemental table S3. Two no-vaccine baselines were constructed: dynamic (CDR and TSR follow the fitted trajectories) and static (both held at their 2015 values). The dynamic baseline was the primary reference because it did not freeze future program performance at a single year’s level. Detection of extrapulmonary disease was held static in both baselines: no EPTB-targeted case-finding initiative comparable to the pulmonary Nikshay and GeneXpert scale-up exists in India, and WHO-reported extrapulmonary burden has been rising over the calibration window while pulmonary burden falls, so a single shared dynamic detection multiplier could not reproduce both disease trends. This asymmetry was the structural reason the two disease types had qualitatively different projected trajectories.

### Vaccine effectiveness and the vaccination schedule

Trial-derived vaccine efficacy estimates were PTB 13·6% and EPTB 42·3% under mITT, and PTB 19·5% and EPTB 50·4% per protocol.^13^ These were combined into effectiveness posteriors by Bayesian evidence synthesis, which is more reflective of real-world vaccine effectiveness rather than direct trial vaccine efficacy. The two efficacies were combined by a power-prior-adjusted conjugate normal–normal update on the log-hazard-ratio scale (per-protocol as prior, discounted by α=0·082 for ∼ 92% participant overlap; mITT as likelihood), giving posterior median effectiveness of PTB 14·0% (95% credible interval [CrI] −19·0 to 37·9), EPTB 42·9% (−5·6 to 69·1) and all-TB 17·2% (−11·7 to 38·6). PTB and EPTB posteriors were drawn jointly via a Gaussian copula (ρ=0·5); the realized correlation across 1,000 draws was 0·50 (online supplemental methods, figure S2, and table S4). Because the posterior credible intervals crossed zero, some sampled draws were negative. The base case truncated these at zero, and the untruncated posterior is reported alongside it as a named sensitivity analysis (online supplemental table S5).

Vaccination was modeled as three discrete, one-time dosing events. Each booster was assumed to confer the same progression-blocking effectiveness as the initial dose, as no revaccination efficacy data exist for VPM1002. The initial dose drew from the never-vaccinated susceptible and latent pools; the two boosters drew from everyone ever vaccinated who is not currently in active disease or on treatment. Dose timing and protection duration were distinct axes, and this analysis held the first constant across both protection durations, so the second can be read cleanly. Under 10-year protection the doses tiled the horizon almost continuously; under 3-year protection each lapsed after three years, leaving seven-year unprotected gaps. The 3-year case was therefore a durability stress-test on a fixed calendar, not a three-yearly revaccination program (online supplemental table S6).

### Health and economic outcomes

DALYs combined years of life lost and years lived with disability, weighted by global burden of disease disability weights,^26 27^ were discounted at 3% per year. Economic analysis was reported as median gross incremental cost-effectiveness ratio (ICER) and net ICER, plus benefit–cost ratio (BCR) with value of a statistical life (VSL) with central value US$80,000. Vaccine cost was assumed at US$0·75 per dose procurement plus US$2·00 administration, US$2·75 per dose event; because the program delivers approximately 1·36 million doses per 1,000,000 targeted, procurement alone is not a sufficient basis for costing a boosted schedule (online supplemental figure S3; unit costs and disability weights in online supplemental tables S7 and S8). Results were benchmarked primarily against the US$500 per DALY opportunity-cost threshold, with 1× and 3× gross-domestic-product-per-capita thresholds (approximately US$2,695 and US$8,084) as secondary benchmarks.^28^ Cases and deaths averted were undiscounted counts, whereas DALYs, costs and benefits were discounted.

### Uncertainty and sensitivity analysis

Uncertainty was propagated through 1,000 Monte Carlo iterations per scenario, jointly sampling the correlated effectiveness posterior. Base-case iterations propagated effectiveness uncertainty only; the 14 calibrated parameters were held fixed, so reported intervals are effectiveness-uncertainty intervals rather than full parameter uncertainty. Counts, DALYs, benefits, costs and the BCR were summarized by mean and 95% uncertainty interval. The gross ICER was summarized by its median, taken over iterations with positive DALYs averted, because DALYs averted appear in the denominator and draws approaching zero produce extreme values that destabilize the mean. Within each protection duration the base seed was matched across the one-, two- and three-dose arms, so each dose’s marginal contribution was a paired difference on identical effectiveness draws. Every result came from a single dose-schedule run against the dynamic baseline; the static baseline was retained only as the calibration comparator (figure 2). An independent replication of the three-dose intervention under 10-year protection, with different seeds, agreed to within 8% (mean cases averted 4,798 against 5,218; mean BCR 17·1 against 18·4); this is Monte Carlo sampling spread and was reported as the practical precision of a 1,000-iteration arm. Variance drivers were ranked by Pearson and partial correlation, and one-way sensitivity analyses covered the VSL grid (US$40,000 to US$416,000),^29 30^ the truncation convention, and the assumed extrapulmonary-share trend (online supplemental figures S4, S5 and tables S5 and S9).

**Figure 2.**
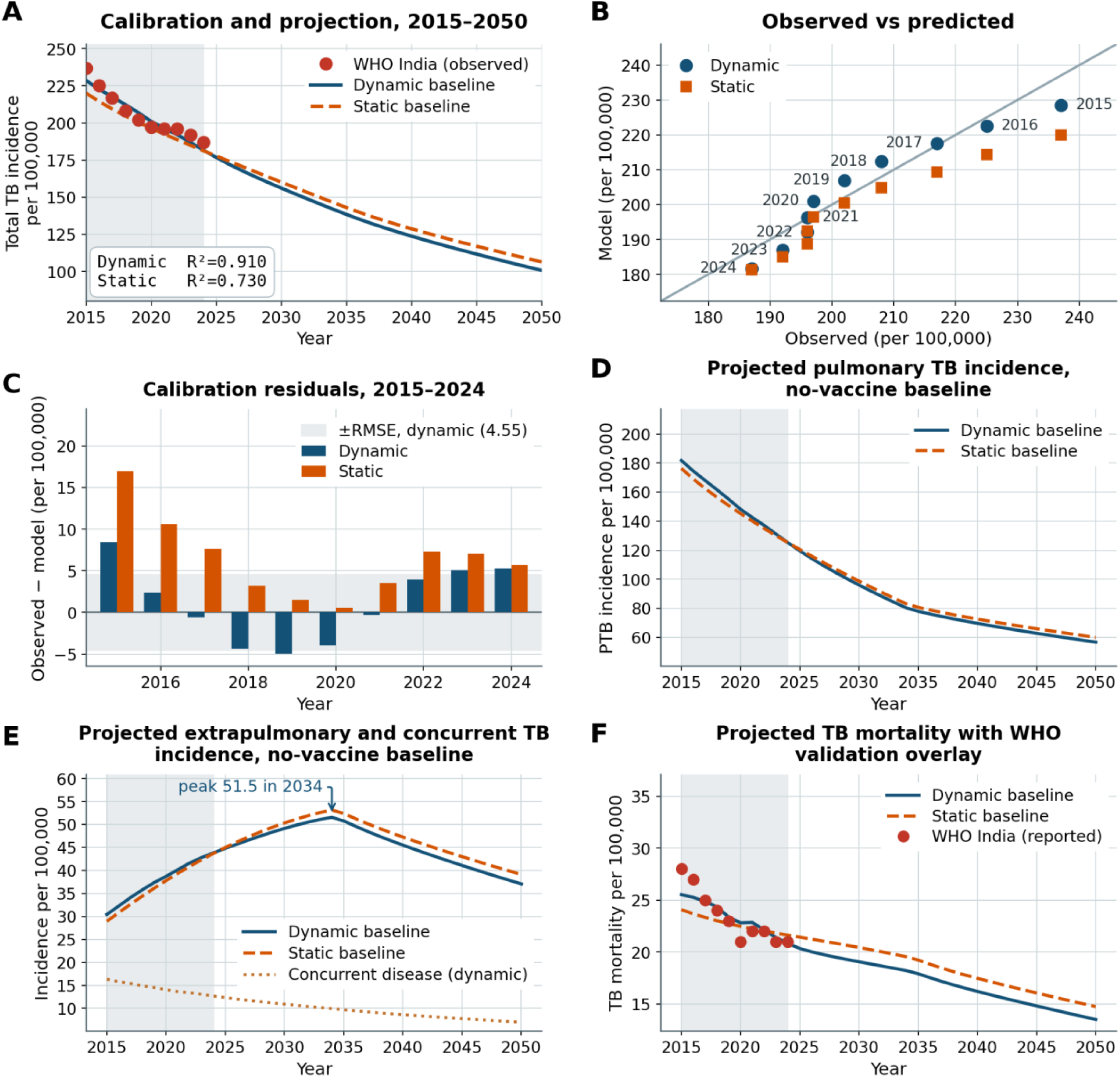
Model calibration and projected no-vaccine baseline trajectories for India, 2015–2050. (A) Annual total TB incidence per 100,000, model against WHO India observed numbers for 2015–2024, with both baselines carried forward to 2050. The shaded band highlights the calibration window; the inset provides each baseline’s R². (B) Observed against predicted total TB incidence, with the line of identity. Each point labelled by year. (C) Calibration residuals for observed minus model total TB incidence, 2015–2024. The shaded band is ±RMSE for the dynamic baseline (4·55 per 100,000). Dynamic residuals change sign across the window: positive in 2015–2016, negative from 2017 to 2021, positive again from 2022 to 2024, indicating no single directional bias. The negative run from 2017–2021 means the model predicted more incidence than observed; only its 2020–2021 portion is consistent with the COVID-era dip in case detection, and the U-shaped pattern also indicates some structural misfit. Static residuals are positive in every year, averaging +6·4 per 100,000, the signature of a baseline that cannot represent program-driven improvements. (D) Projected pulmonary tuberculosis incidence per 100,000 population, India, 2015–2050. (E) Projected extrapulmonary and concurrent tuberculosis incidence per 100,000 population, India, 2015–2050. Extrapulmonary incidence peaked at 51·5 per 100,000 in 2034. (F) Projected tuberculosis mortality per 100,000 population, India, 2015–2050, with WHO-reported estimates overlaid for the 2015-2024 validation window.

### Patient and public involvement

Patients and the public were not involved in the design, conduct, reporting or dissemination plans of this modelling study. This model used only published aggregate epidemiological data and published trial^13^ summary estimates.

## Results

### Model calibration

The dynamic-baseline model reproduced observed TB incidence over the 2015–2024 window (RMSE 4·55 per 100,000; MAPE 1·91%; R²=0·910; figure 2A–B). The static baseline fitted the same ten points markedly worse (RMSE 7·88; MAPE 3·03%; R²=0·730), with residuals positive in all ten years, averaging +6·4 per 100,000 of under-prediction, the signature of a baseline that could not represent program-driven improvement. We choose the dynamic baseline as the primary reference because it holds the vaccine to what it adds over a program (NTEP) that continues on its observed trajectory; a static baseline freezes case detection and treatment success at 2015 levels and so credits the vaccine with burden that continued program improvement would itself have averted. Dynamic residuals changed sign across the window, positive in 2015–2016, negative from 2017 to 2021 (largest −4·95 in 2019) and positive again from 2022 to 2024 (+5·32 by 2024), with the 2020–2021 portion consistent with the COVID-era dip in case detection; the pre-pandemic 2017–2019 deviations and the late positive residuals indicate some residual structural misfit in the shape of the fitted trajectory (figure 2C). CDR followed a shallow logistic curve (54·3% to 92·0%, growth 0·54 per year; R²=0·789) anchored near the observed 2015 value of 53%; TSR rose from a fitted 70·2% in 2015 to 89·1% in 2023 against a 95·0% ceiling (R²=0·903).

### Baseline projections and mortality

Under the dynamic baseline, PTB incidence fell from 181·8 to 125·2 per 100,000 between 2015 and 2024 and continued to 56·7 by 2050 (figure 2D). EPTB incidence rose from 30·4 in 2015 to 43·8 in 2024, peaked at 51·5 in 2034, then declined to 37·1 by 2050, still 22% above its 2015 level (figure 2E). Total incidence fell from 228·5 to 181·7 and then to 100·7 per 100,000. Mortality declined from 25·5 per 100,000 in 2015 to 20·8 in 2024 and 13·5 by 2050, tracking WHO-reported estimates across the validation window (figure 2F). Calibrated mortality fit was R²=0·768. EPTB was the only burden measure that did not decline monotonically; its trajectory followed from the structural asymmetry in detection and was consistent with NTEP notification data showing a rising extrapulmonary proportion (online supplemental table S10).

To assess projections beyond the fitted period, the calibration was repeated using only 2015–2020 data and the resulting parameter set used to forecast 2021–2024 out of sample. Modelled incidence and mortality declined more steeply than subsequently observed, with the divergence increased over the forecast horizon (mean absolute percentage error 8·0% for incidence, 2·5% for mortality). Because vaccine impact is the difference between vaccinated and no-vaccine trajectories, a baseline that declines too quickly narrows rather than widens that gap, so the health and economic benefits reported here are, if anything, conservative.

### The single dose campaign

A single dose given in 2026, with no boosters, was the simplest program the model could evaluate as shown in figure 3. It is reported first because every booster result was measured against it. Under 10-year assumed protection and the dynamic baseline, the vaccine averted 3,306 cases (95% uncertainty interval 91–6,553), 427 deaths (14–802) and 6,751 DALYs (225–12,663) per 1,000,000 targeted, at a mean BCR of 20·7 and a median gross ICER of US$286 per DALY. Under 3-year protection (the trial observation period), the same single dose averted 1,106 cases (39–2,299), 137 deaths (6–270) and 2,454 DALYs (110–4,803), at a BCR of 7·3 and US$779 (table 1 and figure 3). The gap between extrapulmonary disease’s share of averted deaths and its share of averted cases did not depend on the number of doses given. Even with a single dose, EPTB accounted for 60·7% of averted deaths against 44·4% of averted cases under 3-year protection, and 64·4% against 48·2% under 10-year protection.

**Figure 3.**
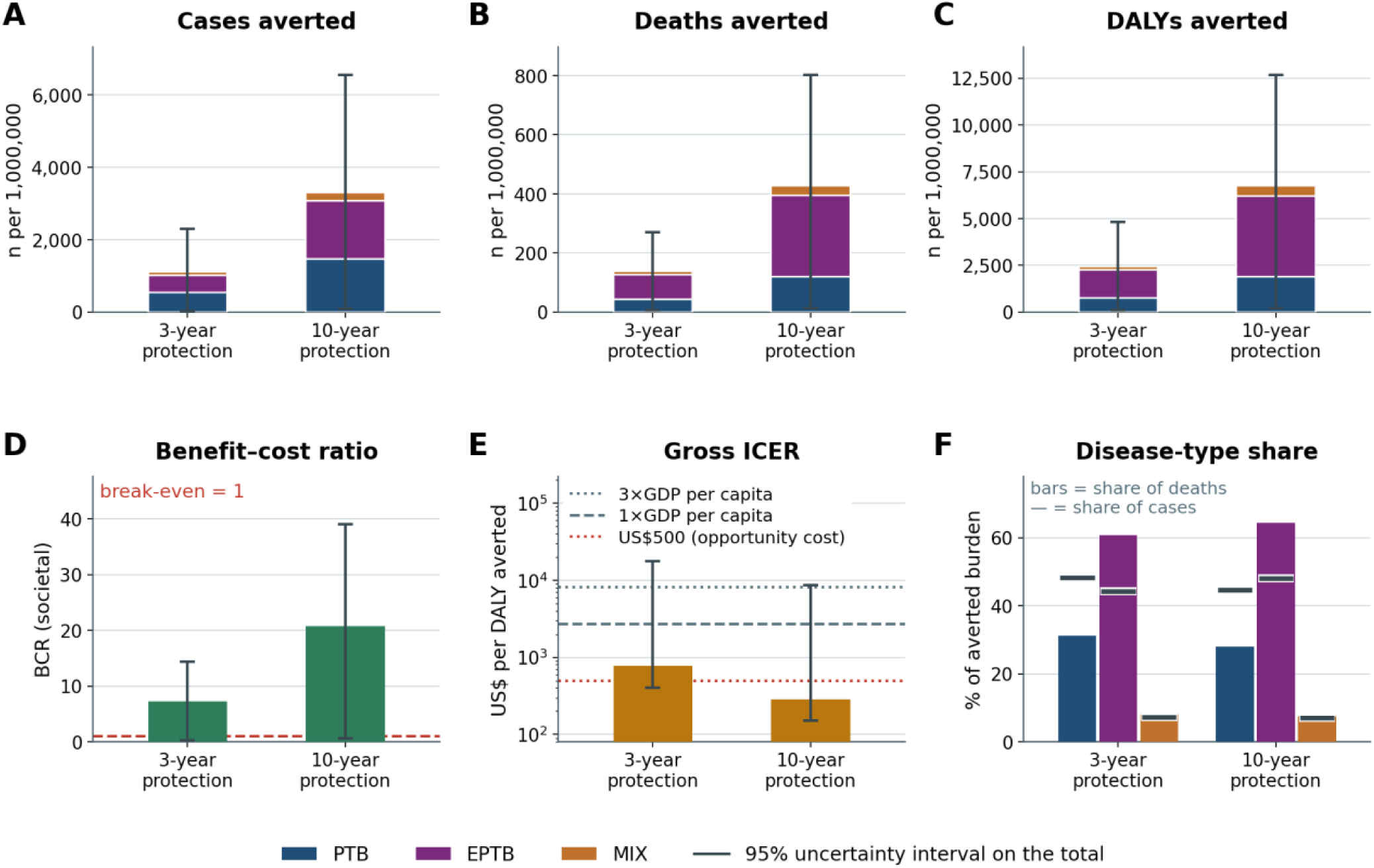
Health impact and cost-effectiveness of the single 2026 dose with no boosters under dynamic baseline, per 1,000,000 population targeted. (A–C) Cases, deaths and DALYs averted, stacked by disease type; error bars are the 95% uncertainty interval on the total. (D) Benefit–cost ratio, societal perspective, value of a statistical life US$80,000. (E) Median gross incremental cost-effectiveness ratio against the US$500-per DALY opportunity-cost threshold and India’s 1× and 3× GDP-per-capita thresholds (∼US$2,695 and US$8,084), log scale. (F) Disease-type share of averted deaths (bars) with the corresponding share of averted cases marked (horizontal ticks). Extrapulmonary disease takes a larger share of averted deaths than of averted cases under both protection durations: 60·7% of deaths against 44·4% of cases at 3 years, and 64·4% against 48·2% at 10 years. Bars and error bars are means with 2·5th–97·5th percentiles over 1,000 iterations, except the ICER, which is the median over iterations with positive DALYs averted. All panels are derived from the dose-schedule run against the dynamic baseline.

**Table 1.** Health impact and cost-effectiveness of each dose schedule, on the fixed 2026 / 2036 / 2046 calendar, dynamic baseline, per 1,000,000 population targeted. Each row is a complete program compared with no vaccination, so the three-dose row includes the effect of all three doses.

| Protection duration | Dose schedule | Disease type | Cases averted (n) | Deaths averted (n) | DALYs averted (n) | BCR | Gross ICER (US\$/DALY) |
| --- | --- | --- | --- | --- | --- | --- | --- |
| 3-year | One dose | PTB | 534 (5–1,317) | 43 (0–107) | 764 (6–1,915) |  |  |
| 3-year | One dose | EPTB | 491 (23–874) | 83 (4–148) | 1,484 (58–2,604) |  |  |
| 3-year | One dose | MIX | 81 (3–162) | 11 (0–22) | 206 (9–405) |  |  |
| 3-year | One dose | All types combined | 1,106 (39–2,299) | 137 (6–270) | 2,454 (110–4,803) | 7.3 (0.3–14.4) | 779 (401–17,599) |
| 3-year | Two doses (10-yearly) | PTB | 747 (8–1,829) | 60 (1–149) | 980 (8–2,451) |  |  |
| 3-year | Two doses (10-yearly) | EPTB | 770 (32–1,356) | 132 (5–231) | 2,097 (82–3,674) |  |  |
| 3-year | Two doses (10-yearly) | MIX | 118 (5–233) | 16 (1–32) | 273 (12–533) |  |  |
| 3-year | Two doses (cumulative) | All types combined | 1,636 (62–3,336) | 208 (10–401) | 3,350 (157–6,460) | 7.3 (0.3–14.1) | 790 (415–17,142) |
| 3-year | Three doses (10-yearly) | PTB | 862 (9–2,114) | 68 (1–170) | 1,043 (8–2,615) |  |  |
| 3-year | Three doses (10-yearly) | EPTB | 960 (38–1,678) | 165 (7–288) | 2,372 (93–4,142) |  |  |
| 3-year | Three doses (10-yearly) | MIX | 139 (6–271) | 19 (1–36) | 293 (13–568) |  |  |
| 3-year | Three doses (cumulative) | All types combined | 1,961 (79–3,965) | 252 (12–480) | 3,708 (179–7,111) | 7.1 (0.3–13.7) | 832 (439–17,562) |
| 10-year | One dose | PTB | 1,476 (17–3,566) | 119 (1–293) | 1,901 (16–4,696) |  |  |
| 10-year | One dose | EPTB | 1,592 (31–2,787) | 275 (5–482) | 4,315 (71–7,547) |  |  |
| 10-year | One dose | MIX | 238 (7–458) | 32 (1–62) | 536 (17–1,015) |  |  |
| 10-year | One dose | All types combined | 3,306 (91–6,553) | 427 (14–802) | 6,751 (225–12,663) | 20.7 (0.7–39.0) | 286 (152–8,653) |
| 10-year | Two doses (10-yearly) | PTB | 2,071 (24–4,978) | 168 (2–411) | 2,447 (21–6,027) |  |  |
| 10-year | Two doses (10-yearly) | EPTB | 2,400 (42–4,191) | 418 (7–729) | 5,906 (93–10,290) |  |  |
| 10-year | Two doses (10-yearly) | MIX | 342 (11–647) | 46 (2–87) | 702 (23–1,320) |  |  |
| 10-year | Two doses (cumulative) | All types combined | 4,814 (138–9,400) | 632 (22–1,165) | 9,055 (311–16,719) | 20.3 (0.7–37.6) | 295 (161–8,724) |
| 10-year | Three doses (10-yearly) | PTB | 2,212 (25–5,325) | 177 (2–434) | 2,518 (21–6,211) |  |  |
| 10-year | Three doses (10-yearly) | EPTB | 2,639 (43–4,595) | 456 (7–793) | 6,221 (98–10,818) |  |  |
| 10-year | Three doses (10-yearly) | MIX | 367 (12–692) | 49 (2–92) | 724 (24–1,359) |  |  |
| 10-year | Three doses (cumulative) | All types combined | 5,218 (153–10,157) | 682 (24–1,255) | 9,463 (328–17,404) | 18.4 (0.6–33.8) | 329 (180–9,638) |

Dose-schedule run, dynamic baseline, 1,000 iterations, fixed 2026/2036/2046 dose calendar. Cases, deaths, DALYs and the benefit–cost ratio are means with the 95% uncertainty interval; the gross ICER is the median with the 95% uncertainty interval over iterations with positive DALYs averted. BCR and ICER are reported once per dose schedule because they are program-level ratios and are not defined by disease type. Under 10-year protection the three-dose row describes the same intervention as the independent replication reported in the main text, estimated in a different Monte Carlo run with different random seeds; the two agree to within 8%.

### Projected impact of the three-dose program

Against the dynamic baseline, the three-dose booster program under 10-year protection averted a mean of 5,218 TB cases (95% uncertainty interval 153–10,157), 682 deaths (24–1,255) and 9,463 DALYs (328–17,404) per 1,000,000 targeted, at a discounted vaccination cost of US$3·13 million (table 1). Under 3-year protection on the same calendar it averted 1,961 cases (79–3,965), 252 deaths (12–480) and 3,708 DALYs (179–7,111), at an almost identical cost of US$3·12 million and an almost identical number of doses (figure 4A–B; online supplemental tables S6 and S7). A 3-year vaccine therefore delivered approximately 38% of the cases and 39% of the DALYs averted by a 10-year vaccine for the same cost and delivery effort, because seven of every ten years between dose events fall outside its protected window. Durability, not schedule intensity, converted doses into averted disease. Cumulative cases and deaths averted stepped up at each dose event and flattened as protection expired (figure 4C). Monte Carlo output distributions across the 1,000 iterations for the three-dose program under 10-year protection and the dynamic baseline were slightly right skewed, so the mean exceeded the median by a small margin in each (online supplemental figure S6).

**Figure 4.**
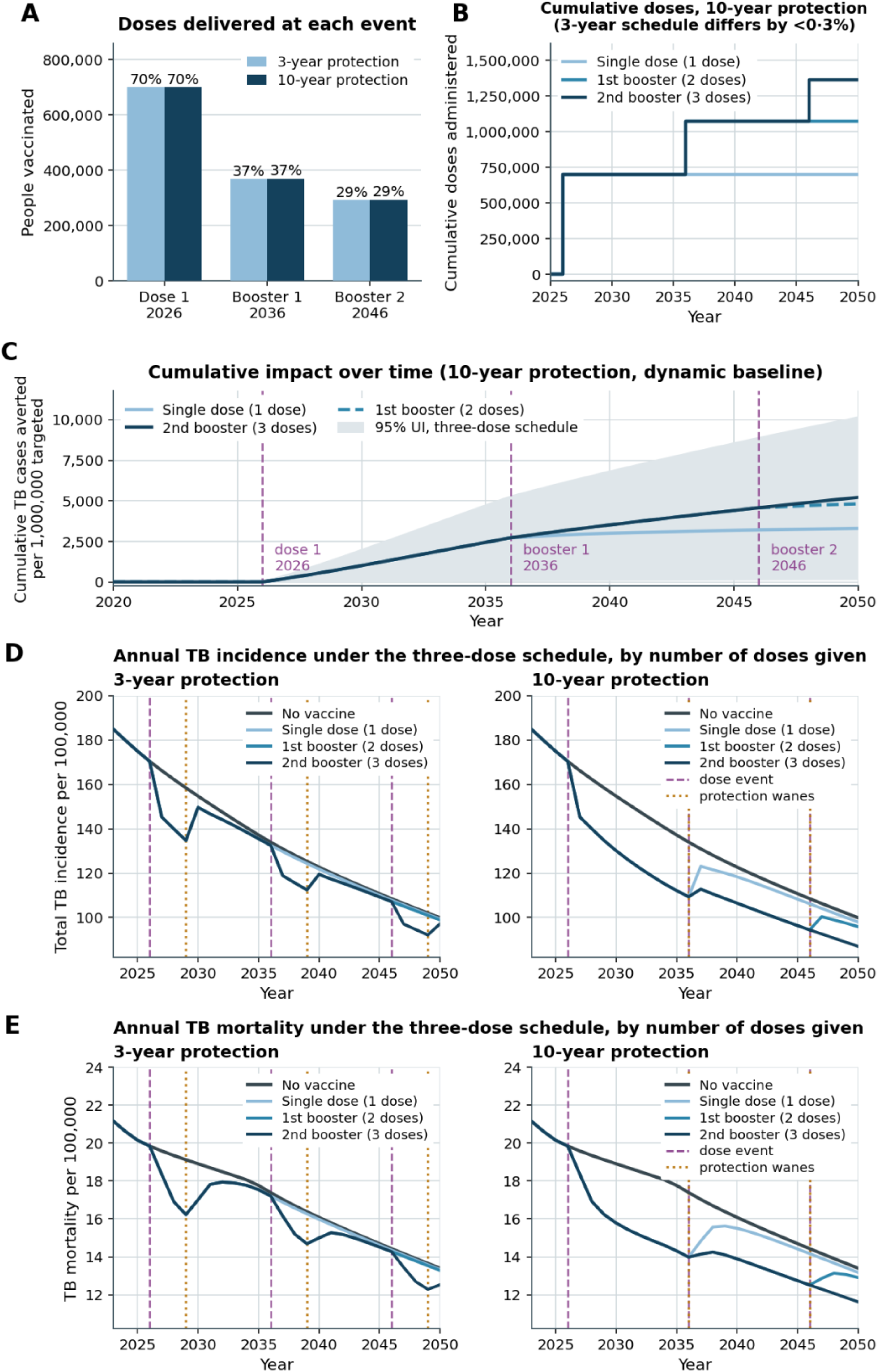
The three-dose schedule: doses delivered, cumulative impact, and annual trajectories by dose count. (A) People vaccinated at each dose event, under 3-year and 10-year protection. The two durations deliver the same doses to the same people: the eligible pool shrinks at each event, so the boosters reach 37% and 29% of the original 1,000,000-person cohort against 70% for the initial dose, irrespective of how long protection lasts. Expressed instead as coverage of the pool eligible at each event, all three doses reach 64·9% (online supplemental table S5). (B) Cumulative doses administered under 10-year protection, by number of doses given. The 3-year schedule differs by less than 0·3% and is not plotted separately. (C) Cumulative TB cases averted over time under 10-year protection, by number of doses given; the shaded band is the 95% uncertainty interval for the three-dose schedule. Vertical lines mark the three dose events. The two-dose and three-dose curves are almost indistinguishable until 2046, which is the second booster’s small marginal contribution seen directly. (D) Annual total TB incidence, and (E) Annual TB mortality, per 100,000 under 3-year protection (left) and 10-year protection (right), by number of doses given, with the no-vaccine baseline for comparison. In D and E, dashed vertical lines mark dose events (2026, 2036, 2046) and dotted lines mark the years protection expires. Under 3-year protection these fall seven years apart, in 2029, 2039 and 2049, leaving unprotected gaps between dose events. Under 10-year protection each dose expires in the year the next is given, so the two sets of markers coincide in 2036 and 2046, and protection is close to continuous. Incidence and mortality fall after each dose event and rebound towards the no-vaccine baseline as protection expires, without exceeding it.

By disease type, extrapulmonary disease contributed the larger share of averted deaths, DALYs and monetized benefit in every dose arm, and the larger share of averted cases in five of six. Across the two three-dose scenarios, EPTB accounted for 49·0–50·6% of averted cases, 65·6–66·9% of averted deaths, 64·0–65·7% of averted DALYs and 62·0–63·8% of monetized benefit, against 42·4–43·9%, 25·9–27·1%, 26·6–28·1% and 28·3–29·9% for PTB (table 1; online supplemental figure S7). Under 10-year protection with all three doses, EPTB accounted for 456 of 682 averted deaths, 6,221 of 9,463 averted DALYs and US$36·6 million of US$57·4 million monetized benefit. The only arm in which pulmonary disease led on averted cases was the single dose under 3-year protection (48·3% against 44·4%); extrapulmonary disease overtook it once a booster was added and led on deaths and DALYs even there. The pattern followed from the product of higher posterior effectiveness, higher case fatality and higher per-case disability weight, which a composite-endpoint model cannot produce. The extrapulmonary share of averted deaths rose with each dose and with longer protection, from 64·4% after the initial dose to 66·9% after two boosters (online supplemental figure S8).

### Marginal value of each dose

Two quantities are reported for each schedule and should not be confused. The cumulative effect was the whole program with that many doses against no vaccination, so the three-dose value already contained the first and second doses (table 1). The marginal effect was what a dose event added on top of the schedule without it, computed as a paired difference on identical effectiveness draws (online supplemental figure S9 and table S11). Because the arms were seed-matched, the decomposition was exact iteration by iteration: 3,306 + 1,508 + 404 reproduced the 5,218 three-dose total under 10-year protection. Successive doses had sharply unequal marginal value (table 1; online supplemental tables S11). Under 10-year protection, the initial 2026 dose alone averted 3,306 cases and 6,751 DALYs at a mean BCR of 20·7 and a median gross ICER of US$286. Adding the 2036 booster raised the total to 4,814 cases (95% uncertainty interval 138 to 9,400) and 9,055 DALYs (311 to 16,719); its paired incremental contribution was 1,508 cases (48 to 2,842) and 2,304 DALYs (82 to 4,191), at an incremental BCR of 19·1 and ICER of US$318. The first booster was therefore nearly as efficient as the initial dose and added 46% more averted cases and 34% more averted DALYs. The 2046 booster raised the total to 5,218 cases (153 to 10,157) and 9,463 DALYs (328 to 17,404), but contributed only 404 cases (14 to 742) and 408 DALYs (10 to 721), at a BCR of 6·8 and ICER of US$1,051. Two plausible mechanisms explained the drop: the number vaccinated shrank at each event (700,000, then approximately 370,000, then approximately 294,000, from eligible pools of approximately 1,076,000, 569,000 and 451,000 individuals, respectively), and only four of the second booster’s ten protected years fell inside the 2050 horizon.

Under 3-year protection assumption, the pattern was similar but less extreme, because each dose’s protection was fully realized inside the horizon: the initial dose averted 1,106 cases, the first booster added 529 (24 to 1,039) at an incremental BCR of 7·2, and the second added 325 (16 to 618) at 5·9. In both scenarios the annual trajectories showed the expected sawtooth, incidence falling sharply after each dose event and rebounding towards the no-vaccine baseline as protection expired, without ever exceeding it (figure 4D–E). Absolute impact varied 4·7-fold across the six dose-by-duration arms; normalizing each to its protected person-time compressed that range to 439 to 527 averted cases per million protected person-years, so the differences between arms reflect exposure to protection rather than differing efficiency.

### Cost-effectiveness

Mean BCR (societal perspective, VSL US$80,000) was 18·4 (95% uncertainty interval 0·6 to 33·8) for the three-dose program under 10-year protection and 7·1 (0·3 to 13·7) under 3-year protection; both exceeded break-even, although the lower bound fell below 1 in every arm (table 1; online supplemental figure S10). Median BCR stayed above break-even at every VSL from US$40,000 to US$416,000, the lowest anywhere on the grid being 4·7 (online supplemental table S12). Median gross ICER was US$329 per DALY averted under 10-year protection and US$832 under 3-year protection, ranging US$286–1,191 across individual and incremental dose comparisons; all fall below India’s 1× GDP-per-capita threshold of approximately US$2,695, the least favorable being less than half of it. This GDP comparison is secondary to the US$500 per DALY opportunity-cost benchmark, at which the probability of cost-effectiveness was 92·3% under 10-year and 67·1% under 3-year protection. Net cost was US$2·42 million under 10-year protection, so once averted treatment and hospitalization costs were counted the program was cost-saving on average from the health-system perspective and therefore dominant (more DALYs averted at lower net cost; dominant in 81·5% of iterations); under 3-year protection it was +US$0·97 million, a median net ICER of US$252 per DALY averted. The mean gross ICER was not a stable statistic in any scenario; the median and the acceptability curves are the appropriate primary outputs (figure 5A).

**Figure 5.**
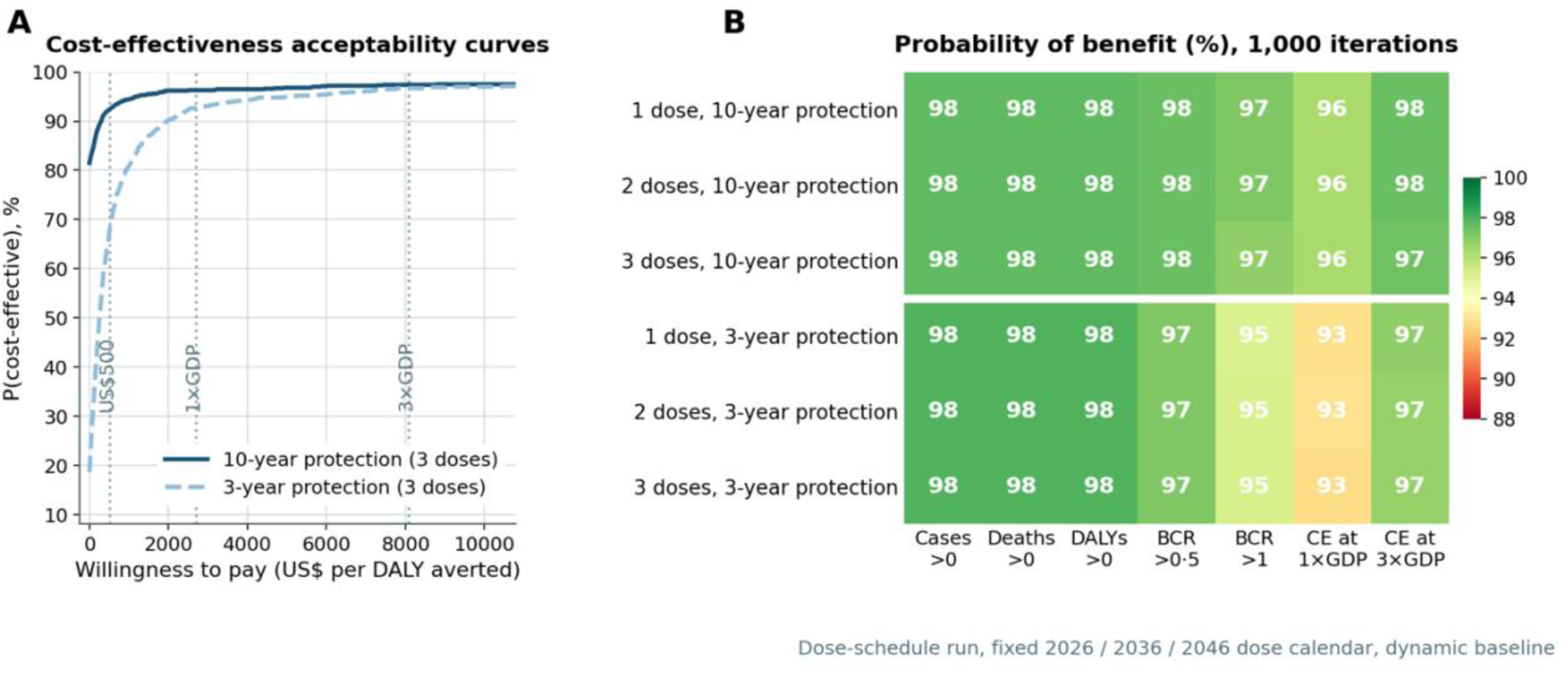
Cost-effectiveness acceptability curves and probability of benefit. (A) Cost-effectiveness acceptability curves for the two full three-dose programs against willingness to pay, with the US$500, 1× GDP and 3× GDP thresholds marked. Note the truncated y-axis. The two durations separate sharply below 1× GDP per capita, where the 10-year program is already above 93% and the 3-year program rises from about 70%, and converge near 97% above it. (B) Probability, across 1,000 iterations, that averted cases, deaths and DALYs exceed zero, that the benefit–cost ratio exceeds 0·5 and 1·0, and that the program is cost-effective at 1× and 3× GDP per capita, for each of the six dose-schedule arms: three dose counts at each of two protection durations, seed-matched within a duration. Rows above the white rule are the 10-year protection arms; rows below are the 3-year arms. Values are near-identical across dose counts within a duration, because the probability of benefit depends on the sign of the sampled effectiveness draw rather than on how many doses are given. These probabilities have a ceiling set by the effectiveness-truncation convention: in 2·1–2·3% of iterations both effectiveness draws are truncated to zero, so no vaccine effect is modeled. Dose-schedule run, fixed 2026/2036/2046 dose calendar, dynamic baseline.

### Probability of benefit and drivers of uncertainty

The probability of a positive health effect was at least 97·7% for cases, deaths and DALYs in every scenario and dose arm (figure 5B). This was bounded from above by the truncation convention: in 2·1–2·3% of iterations both effectiveness draws were truncated to zero, so no vaccine effect was modeled. Without truncation, 7·2% of iterations were net-negative for cases averted, and truncation inflated mean cases averted by 5·9% and mean DALYs averted by 5·0% (online supplemental table S5). Differences between cases, deaths and DALYs within a scenario were smaller than that zero-effect group and should not be interpreted as real differences. The probability that BCR exceeded 1 was 95·1–97·1%, and of cost-effectiveness at 3× GDP per capita 96·7–97·5% (online supplemental table S13). Vaccine effectiveness was the only resampled input in the base case, so the variance decomposition ranked its two components against each other rather than against the fixed calibrated, epidemiological and cost inputs. Across the six dose-by-duration arms the pulmonary correlation with BCR ranged 0·85–0·91 and the extrapulmonary 0·82–0·88. The bands overlapped and no ranking is claimed. Extrapulmonary disease dominated the level of averted mortality and disability, while neither endpoint dominated the variance of the benefit–cost ratio, because the two draws were correlated by construction.

### Demographic distribution of impact

Impact concentrated in working-age adults. Under 10-year protection with all three doses, the 31–55-year band accounted for 39·9% of combined pulmonary and extrapulmonary cases averted, the 15–30-year band 31·7%, the 56–80-year band 19·4% and the 6–14-year band 9·0% (online supplemental figure S11, and table S14). Impact concentrated in the HIV-negative majority (96·8%), the normal-body-mass-index stratum (50·5%) and the middle-socioeconomic stratum (43·2%). Because effectiveness was not modeled as stratum-varying, these patterns reflected where burden and uptake fell rather than differential biological effect; the low-socioeconomic stratum showed fewer absolute cases averted than the middle stratum despite higher disease burden, because modeled uptake was lower (0·85 against 1·00). The 6–14-year band, the age group the trial uniquely enrolled, was 20% of the modeled population and received the highest uptake multiplier, yet contributed under a tenth of averted cases, because Indian TB burden is concentrated in adults.

## Discussion

This is, to our knowledge, among the first TB transmission models to track pulmonary and extrapulmonary disease as separate compartments with independently parameterized natural history, separately propagated effectiveness distributions, separately reported health and economic outcomes, and an explicit multi-dose schedule in which each dose’s marginal contribution can be estimated rather than inferred. The model uncovered two main findings. The first is a dissociation between where the vaccine averts cases versus where it averts harm. Extrapulmonary disease contributes the larger share of averted deaths, DALYs and monetized benefit in every scenario and at every dose count, and by a far wider margin than it contributes to averted cases: roughly two-thirds against roughly half. This cannot be derived by decomposing a composite-endpoint model after the fact because it depends on the product of type-specific effectiveness, case fatality and disability weight, which do not cancel when aggregated. It requires that the two disease types be tracked separately from the force of infection through to health-economic valuation to additionally capture their different transmission and detection dynamics. The mortality and DALY dissociation is large, roughly two to one over pulmonary disease, and holds across every scenario and dose count. The shares of averted cases are much closer together, so the case-count ranking is the less robust of the two comparisons. The second is that the marginal value of boosting is highly unequal across doses. One booster a decade after the initial dose adds 46% more averted cases and 34% more DALYs, at close to the same economic efficiency. A second booster dose adds only 8% more cases averted and 4.5% more DALYs at roughly a third of that efficiency. Some of this is a real feature of a waning vaccine deployed into a shrinking eligible pool, and some is an artefact of the horizon, since six of the second booster’s ten protected years fall after 2050. The distinction matters for policy: the case for a single decadal booster is robust here, while the case for a second is horizon-sensitive and should be re-examined over a longer projection window.

The disproportionate extrapulmonary contribution to averted mortality and DALYs is consistent with VPM1002’s mechanism of action and with the epidemiology of extrapulmonary disease. CD8 T-cell and trained-innate responses have been proposed as the downstream mechanism; if that holds, protection would not depend on prior *M. tuberculosis* sensitization. That matters because EPTB disproportionately affects HIV-positive, malnourished, and immunocompromised individuals^6 31 32^ who are frequently IGRA-negative as a consequence of immune dysfunction rather than absence of infection. Extrapulmonary protection from this platform is therefore mechanistically plausible, though the molecular mechanisms have not been elucidated for this vaccine in humans. Diagnostic constraints compound the problem: with confirmation achieved in only 16% of EPTB cases, confirmed-endpoint analyses may attenuate the signal, though the magnitude and direction of any ascertainment effect are uncertain. Caution in interpreting secondary endpoints is warranted given the primary endpoint did not meet significance, and larger studies would strengthen confidence. For policy purposes, however, ignoring the available extrapulmonary signal carries its own risk of misallocation, and the present model quantifies what is lost by doing so.

The broader TB vaccine modelling literature has been anchored to M72/AS01ᴇ, a subunit vaccine with a different mechanism and a target population enriched for IGRA-positive adults. Clark and colleagues^16 17^ and Sumner and colleagues^33^ conducted cost-effectiveness analyses in that framework without disaggregating pulmonary and extrapulmonary outcomes. Hatherill and colleagues^34^ argued that young adolescents aged 6 to 14 years should be a priority population for novel TB vaccines but concluded the evidence was insufficient to support their inclusion in current policy, drawing exclusively on M72/AS01ᴇ-derived modelling; the PreVenTB Phase III trial^13^ modelled here enrolled household contacts from age 6 upward and is, to our knowledge the only Phase III TB vaccine trial to have done so. Our model includes the 6 to 14-year band as one of four stratified age bands and provides a trial-grounded quantification for this age group. It also shows that this band accounts for under a tenth of averted cases in absolute terms. The policy case for including young adolescents therefore rests on trial evidence, on lifetime benefit from early vaccination, and on equity, rather than on this band carrying the largest share of near-term averted burden.

A concurrent analysis by Clark and colleagues modeled VPM1002 across 79 low-and-middle income countries (LMICs).^35^ They estimated that introduction of VPM1002 might avert 6·7 million symptomatic TB episodes and 0·7 million deaths by 2050, and that it might be cost-effective in 15 of 79 countries from the health-system perspective. No head-to-head comparison with the present analysis is possible, and none is attempted. The two models differ in scope (79 countries against one country where the trial was conducted), calibration method (history matching with emulation against differential evolution), delivery design (routine annual vaccination of the age-six cohort plus 2 campaigns against three discrete campaign doses), baseline (non-vaccine intervention held at 2023 levels against a fitted dynamic program trajectory) and, most consequentially, endpoint: Clark and colleagues used the composite all-TB estimate and reported that the post-hoc extrapulmonary effect was not evaluated there. Thus, the two studies are complementary. Where a multi-country analysis establishes the scale of the global opportunity, this model used India, a LMIC where the trial was conducted, as a case study, so a single national program can see what the vaccine does to each disease type, in each sociodemographic stratum, at each dose event, and decide whether and when to boost. Three modelling structures allow for this distinction: separate pulmonary and extrapulmonary posteriors; vaccination history preserved through a full disease episode, so people infected after vaccination remain eligible for later doses; and dated dose events whose marginal contributions are separately estimable. Two further differences are valuation choices rather than capabilities. We monetized averted mortality with a value of a statistical life, and cost every dose event at procurement plus administration rather than per course. These choices move our benefit–cost ratios relative to theirs (valuation with a value of a statistical life raises them; costing every dose event lowers them) and should be read as such, rather than as evidence of a better vaccine. Conversely, their multi-country calibration propagates parameter uncertainty that our effectiveness-only interval does not.

Other analyses from the same modelling group point in the same direction. Multi-country analyses across 105 low- and middle-income countries,^36^ and the macroeconomic valuations,^20^ establish the scale of the opportunity but treat tuberculosis as one disease and a vaccine as one course, so neither can say where within the disease spectrum the benefit falls. Recent Indian work shows projected impact varies roughly three-fold with the disease stage a vaccine is assumed to act against,^37^ and the field is debating whether asymptomatic disease should be a licensure endpoint at all.^38^ These studies highlight the caveats of aggregating TB disease types; the model we describe resolves disease state and disease type instead of aggregating them so that the contribution of extrapulmonary disease is quantified rather than absorbed into a composite endpoint.

The dynamic baseline reveals a dimension of VPM1002’s value that static models cannot capture. Under NTEP-continuation, PTB incidence declines monotonically as program detection improves, but EPTB incidence continues to rise into the mid-2030s before trending downwards, the only burden measure in this model that moves against the program trend. This reflects a structural asymmetry in what the program can reach: GeneXpert, cough-based case-finding and Directly Observed Treatment, Short-course adherence support are predominantly pulmonary-directed, while extrapulmonary diagnosis depends on clinical suspicion, imaging and biopsy, none of which current NTEP activity meaningfully scales. The same asymmetry applies to TPT; its protection erodes within about a year in high burden settings, driven by reinfection against which it offers no defense.^7^ In our model, VPM1002’s protection is assumed to lower the rate at which latent infection progresses to disease, whether vaccination occurs before or after infection, for as long as protection lasts. The incremental benefit VPM1002 adds over the existing program is therefore proportionally larger for extrapulmonary disease. For pulmonary disease the vaccine supplements a trajectory already improving; for extrapulmonary disease it acts where the program has no equivalent lever. This does not diminish the meaningful pulmonary benefit, which still generates over two-fifths of averted cases and substantial averted mortality in absolute terms. Rather, it identifies where the vaccine is most complementary to the existing program: precisely where detection-based tools are least effective.

This study has limitations. Uncertainty intervals propagate vaccine-effectiveness uncertainty only, not uncertainty in the 14 calibrated parameters, the calibration targets, the cost and disability inputs, or the assumed extrapulmonary-share trend, and are correspondingly narrower than full parameter uncertainty would imply. The rising extrapulmonary share is an imposed structural assumption, selected as the best-fitting of four tested values rather than jointly estimated. Because the posterior credible intervals cross zero, some sampled effectiveness draws were negative, and the base case truncated these at zero. A negative draw implies a vaccine that increases TB risk, for which no mechanism is known in a BCG-derived platform: no serious adverse events attributable to VPM1002 were reported in the Phase III trials,^13 39^ the vaccine was at least as safe as BCG, and a century of BCG use provides no evidence that vaccination causes tuberculosis. The negative tail therefore reflects sampling uncertainty in a trial powered to detect 50% efficacy against a substantially smaller observed effect. Truncation remains a reporting choice, inflating mean averted cases by 5·9% and DALYs by 5·0% and setting the ceiling on the reported probability of benefit; both conventions are reported and the vaccine remained beneficial under either. The transmission system is deterministic, so within-stratum stochastic effects are not represented, which may matter most in the HIV-positive stratum (approximately 3% of the population). Relapse from the on-treatment compartment is modeled as a flow into active disease without a matched subtraction from the treatment pool, because individuals cannot be tracked in a compartmental representation and the outflow terms are specified to avoid double-counting people rather than episodes; this is a recognized limitation of compartmental models generally. Effectiveness inputs were not stratified by age, HIV status, body-mass index or socioeconomic status, so the demographic distribution of impact reflects burden and uptake rather than differential biological effect. Stratum-specific efficacy estimates of this kind are not reported by TB vaccine trials, so no model can currently incorporate them; the constraint is in the evidence base rather than in the model structure, and subgroup estimates from a trial of this size are likely to be too imprecise to propagate usefully. The 2046 booster’s value is truncated by the 2050 horizon under 10-year protection, and the model contains no age stratum below 6 years, so it cannot address infant vaccination or BCG replacement. A routine program that also vaccinated cohorts reaching age 6 later would avert additional cases not captured here.

## Conclusions

In this case study, VPM1002, a candidate TB vaccine, yielded favorable economic outcomes on the central estimates across the modelled protection durations, dose counts and tested values of a statistical life; the probability of a benefit–cost ratio above 1 is 95·1–97·1%, not certainty. The mean societal benefit–cost ratio of the three-dose program was 7·1–18·4 at a value of a statistical life of US$80,000, driven largely by the valuation of averted deaths. Per 1,000,000 targeted, between 252 and 682 deaths and between 3,708 and 9,463 DALYs are averted, with 10-year protection delivering roughly two and a half times the health gain of 3-year protection at the same delivery cost, because durability rather than dose count determines how much of the horizon is protected. A single booster a decade after the initial dose increases averted cases by 46% at close to the efficiency of the initial dose; a second adds substantially less and is horizon-sensitive. The vaccine’s largest point estimate of efficacy is against extrapulmonary TB, although the confidence interval remains compatible with no effect. Extrapulmonary TB is hardest to diagnose, most likely to cause death and lasting disability, and most concentrated in people living with HIV or malnutrition, and it accounts for roughly two-thirds of averted mortality and DALY burden against roughly half of averted cases. Working-age adults benefit most in absolute terms. The schedule evaluated here is an adolescent and adult booster program and is not a substitute for neonatal BCG vaccination. The framework is extensible to other settings, schedules and candidates subject to recalibration. These findings are conditional on the calibration, the assumed extrapulmonary trajectory, effectiveness assumptions, economic inputs and the fixed horizon.

## Data Availability

No individual-level participant data were used for this modelling study. Epidemiological data are available from the WHO Global Tuberculosis Report and India NTEP. The model's full parameter values, calibration targets, epidemiological data sources, and health-economic inputs are reported in the supplementary appendix, together with the code version, calibration cache checksum and run settings that produced every reported number (online supplemental table S15). The differential equation system and implementation code are proprietary to Aseesa Inc. and are not publicly released. The calibrated model is accessible as a runnable interface at Hugging Face (URL available from the corresponding author upon publication), where all reported scenarios can be reproduced and additional analyses run.

## Author contributions

AB and AC conceived the model design. MR wrote all the ODEs for the model with help from VN and AC. MR, VN and VS collected all the input parameters. MR, VS, VN, AU and AC tested the various iterations of the model. MR, VS, AU and AC generated the outputs. MR, AC, AU, VN, and AB contributed to data analysis and interpretation. MR, AU and AB contributed to the preparation of the original draft. All authors had full access to all the data and had final responsibility for the decision to submit for publication.

## Competing interests

MR and VN are affiliated with Aseesa Inc. AB is the founder and an employee of Aseesa Inc. AC is a co-founder of Aseesa Inc.

## Funding

In part funded by Serum Life Science Europe GmbH. The funder had no role in model design, data analysis, interpretation, or writing.

## Patient consent for publication

Not required.

## Ethics approval

Not required. This modelling study used only publicly available aggregate data and published trial summary estimates; no individual-level participant data were accessed.

## Acknowledgements

Claude Code (Anthropic [Opus 5.5]; Aseesa’s institutional license) was used to assist with coding. All code and model outputs were independently checked and verified by the authors (RM, VN, VM, AU, and AC). We gratefully acknowledge the data sharing and support provided by Serum Life Science Europe GmbH. We thank Prof. Richard White and Dr. Rebecca Clark (London School of Hygiene and Tropical Medicine) for generously reviewing this modelling work and providing constructive methodological feedback that strengthened the calibration and baseline projection framework. We also thank Saumya Mehta, and members of the AI-ML team at Aseesa Inc. for help with cross-checking ODEs, data gathering, and verification. We dedicate this work to all the individuals who selflessly participated in TB trials to make this work possible.

## Notes

### Competing Interest Statement

The authors declare the following competing interests: MR and VN are affiliated with Aseesa Inc.; AB is the founder and an employee of Aseesa Inc.; AC is a co-founder of Aseesa Inc. The study was in part funded by Serum Life Science Europe GmbH, which had no role in model design, data analysis, interpretation, or writing. VS declares no competing interests.

### Author Declarations

No individual-level participant data were used for this modelling study. Vaccine efficacy data were derived from: Singh M, Joshi S, Vohra V, et al. Efficacy and safety of VPM1002 and Immuvac in preventing tuberculosis: phase 3 randomised clinical trial (PreVenTB trial). BMJ 2026;393:e085716. Epidemiological data are available from the WHO Global Tuberculosis Report and India NTEP.

### Summary of Updates

The revised paper now presents a modeling method, with VPM1002 in India as a case study, and the title has changed accordingly. The method tracks pulmonary, extrapulmonary and concurrent tuberculosis separately, stratifies by nutritional and socioeconomic status, uses a dynamic programmatic baseline, and estimates what each vaccine dose adds. The population compartments increased to 1,152 (16 states across 72 strata) for a total of 2,160 state variables. New features are a subclinical stage for pulmonary and concurrent disease, compartments for people whose protection has waned, and tracking of vaccination history through disease, treatment and recovery, so that people infected after vaccination remain eligible for later doses. Three discrete booster dose events (2026, 2036 and 2046) under assumed protection of 3 or 10 years are now clearly modeled. Paired Monte Carlo draws across one-, two- and three-dose schedules allow each dose's marginal contribution to be estimated. We added a sensitivity analysis on truncating negative effectiveness draws at zero, an independent replication run, and explicit statements of what the uncertainty intervals include and exclude. Extrapulmonary disease accounted for about two-thirds of averted deaths and DALYs but about half of averted cases. A booster a decade after the first dose is nearly as economically efficient as the first dose; a second booster adds much less. We state explicitly that the trial's confidence intervals include no effect, that results are conditional projections and not evidence that the vaccine works, and that the schedule is an adolescent and adult booster program, not a replacement for infant BCG vaccination. The figures, tables and supplementary materials have been revised to reflect the updated output.

