## Supplemental Methods and Figures S1-S17 for "A disaggregated dynamic model of pulmonary and extrapulmonary tuberculosis for vaccine and booster-schedule evaluation: VPM1002 in India as a case study"

### Online Supplementary Methods and Tables

#### 1. Force of infection

Transmission was age-assortative and frequency-dependent. The force of infection on age band a was $\lambda_a = \beta_{eff} \cdot \Sigma_{a'} C[a,a'] \cdot I_{eff,a'} / N_{a'}$, where $\beta_{eff}$ is the transmission coefficient multiplied by a time-varying scale factor, C is a four-band aggregation of the Prem et al.[1] India synthetic contact matrix, $N_{a'}$ is the total population in age band a′, summed across all sixteen population compartments, vaccinated and waned included, since omitting any of them would inflate λ by shrinking its own denominator, and the effective infectious population is $I_{eff} = I_{PTB} + \rho_{EPTB} \cdot I_{EPTB} + \rho_{MIX} \cdot I_{MIX} + m_{sub} \cdot I_{PTB\_SUB} + m_{sub} \cdot \rho_{MIX} \cdot I_{MIX\_SUB}$. The relative-infectiousness weights were $\rho_{EPTB} = 0{\cdot}30$ and $\rho_{MIX} = 0{\cdot}65$: extrapulmonary and concurrent disease therefore contributed to transmission at reduced weight, not zero. Subclinical disease contributed at $m_{sub}$ times its disease type's own weight, where $m_{sub}$ is a calibrated parameter. A socioeconomic mixing factor was applied multiplicatively to $\lambda_a$, and susceptibility multipliers of 3·5 for HIV-positive status and 1·6 / 1·00 / 0·85 for low / normal / high BMI scaled the stratum-specific hazard.

#### 2. Demography

The population's age structure was initialized using 2011 Census proportions (20% / 30% / 35% / 15% across the four age bands) and evolved dynamically over the simulation horizon via continuous exponential-dwell-time aging with rates 1/9, 1/16, 1/25 and 1/25 per year, age-targeted births, and a uniform background (non-TB-related) mortality rate. In the population compartment, births accrued at 0·025 per year applied to the total population, a rate proportional to population, not a fixed number of births per year. All newborns entered the susceptible compartment of the youngest age band, HIV-negative, distributed across the BMI and socioeconomic strata by the same proportions used to initialize the model. Background non-TB mortality removed individuals from every population compartment at 0·012 per year. Aging was applied to the sixteen population compartments and to the seven vaccination-history counters, whose people age alongside their parent compartments, but not to the seven cumulative counters, which are running totals rather than populations. Individuals aging out of the oldest band left the model, so the net growth rate was births less background mortality, TB deaths and that exit. No negative compartment values and no missing values across all 2,160 compartments and every year of the simulation, with the total population growing from 1,000,000 at the start of burn-in to 1,214,583 in 2050 under the dynamic baseline, a mean growth rate of approximately +0·49% per year were verified numerically for the run reported here.

#### 3. Designing the three-dose program

Vaccination was not modeled as a continuous coverage ramp, but as three discrete, one-time dosing events at the fixed calendar years $t \in \{2026, 2036, 2046\}$, the same calendar at both protection durations. At each event a fraction of the eligible population was moved instantaneously into the protected compartments (V, E_fast_V, E_slow_V), weighted by the stratum-specific uptake multipliers $u_{v,age}$ = (0·90, 0·70, 0·60, 0·50) and $u_{v,SES}$ = (0·85, 1·00, 1·08), applied uniformly across HIV and BMI strata. The eligible pool differed by dose number. The initial dose drew only from the never-vaccinated pools (S, E_fast, E_slow). Doses two and three drew from everyone ever vaccinated who were not currently in active disease or on treatment, i.e., V + E_fast_V + E_slow_V + S_W + E_fast_W + E_slow_W; people in active disease or on treatment became eligible again once R_VHIST resolved into S_W.At each event a fraction of the eligible population was moved instantaneously into the protected compartments (V, E_fast_V, E_slow_V),

The coverage parameter was solved numerically, rather than assumed, so that the initial dose vaccinates exactly 700,000 people, i.e., 70% of the original 1,000,000-person cohort. The solved value was 0·650675. Setting a nominal 70% instead would have vaccinated approximately 753,000 people, because sixteen years of births would grow the eligible pool from 1,000,000 at the 2010 simulation start to approximately 1,076,000 by the 2026 dose event; 0·650675 of that larger pool is 700,000 of the original cohort. Vaccine protection was assumed to be progression-blocking only. Infection-blocking efficacy $\varepsilon_{inf}$ was set to zero on the trial evidence; the disease-type-specific progression-blocking parameters $\varepsilon_{prog,PTB}$, $\varepsilon_{prog,EPTB}$ and ε_prog,MIX defaulted to zero in the parameter class and were overwritten once per Monte Carlo iteration by that iteration's sampled posterior effectiveness draw. Protection expired as a discrete event at the dose year plus the protection duration, at which point the entire protected cohort was moved to the waned pools.

#### 4. Cost and benefit

Vaccination cost was the number of dose events multiplied by US$0·75 procurement plus US$2·00 administration, discounted at 3% per year from a 2026 base year. Averted treatment and hospitalization costs were the discounted downstream savings; averted productivity losses and the value-of-statistical-life valuation of averted deaths were added to obtain the total societal benefit. The benefit–cost ratio was the total societal benefit divided by the vaccination cost. Net cost was the vaccination cost less the treatment and hospitalization savings; negative values are net savings. The gross incremental cost-effectiveness ratio was the vaccination cost divided by discounted DALYs averted, and the net ratio used net cost in the numerator; both are undefined when DALYs averted are not positive. Incremental net monetary benefit at willingness to pay $\lambda$ is $\lambda \times$ DALYs averted − net cost, and the cost-effectiveness acceptability curve reported the probability that this was positive across a grid from zero to four times GDP per capita.

#### 5. Calibration of the dynamic baseline: fit achieved

The no-vaccine baseline used throughout allowed case detection and treatment success to continue along their fitted logistic trajectories. Fitted against the ten observed years it gave $R^2$=0·910 with RMSE 4·55 per 100,000 and MAPE 1·91%, against $R^2$=0·730 and RMSE 7·88 for the static comparator in which detection and treatment success were held at their 2015 values (table S10; figure 2A–B of the main document). The residual structure was more informative than the summary statistics: residuals changed sign across the window: positive in 2015–2016, negative from 2017 to 2021, positive again from 2022 to 2024, so there was no single directional bias; the negative run from 2017 to 2021, in which the model predicted more incidence than was observed, is consistent with the COVID-era dip in case detection only for 2020–2021, and the U-shaped pattern also indicates some structural misfit (figure 2C). Every number reported in this paper comes from the single run pinned in table S15.

#### 6. Cumulative and marginal effects

Each dose-count arm was a complete program: the one-dose arm gave a single dose in 2026, the two-dose arm gave 2026 and 2036, and the three-dose arm gave 2026, 2036 and 2046, and each was compared against the same no-vaccine scenario. The values reported for an arm are therefore cumulative; the three-dose arm's 5,218 averted cases under 10-year protection include the contribution of all three doses. The marginal effect of a single dose event was obtained by differencing two adjacent arms within the same iteration. The cumulative series was also monotone non-decreasing in dose count for every outcome at both protection durations, and the by-disease-type components summed to the arm total in every arm. The cumulative-over-time curves in figure 4C and in figure S7 reached the endpoint totals reported in table 1 of the manuscript. The two are the same quantity computed by two ways: a running sum of annual averted cases versus a single difference of cumulative counters, so the identity holds by construction, and the check confirms that the implementation preserves it. Normalizing each arm's averted burden to the person-time actually spent under protection, defined as the number vaccinated at each dose event multiplied by the protected years falling inside the analytic horizon to 2050, compressed a 4·7-fold range of absolute impacts into 439 to 527 averted cases per million protected person-years, a 1·20-fold spread. The differences between arms therefore reflect how much protected person-time each schedule delivers rather than any change in how efficiently protection converts into averted disease. The residual gradient was downward across successive doses and across durations, in the expected direction: a dose given later protected a smaller surviving and eligible cohort, during years in which the no-vaccine baseline incidence had already declined.

**Online supplemental table S1.** Model structure: 30 states across 72 population strata

16 population compartments + 7 cumulative counters + 7 vaccination-history counters = 30 states; 30 × 72 strata = 2,160-state variables.

| Symbol | Description | Type |
|---|---|---|
| S | Susceptible, never vaccinated | Population |
| V | Vaccinated and currently protected | Population |
| E_fast, E_slow | Latent infection (fast and slow pathway), never vaccinated | Population |
| E_fast_V, E_slow_V | Latent infection, vaccinated and currently protected | Population |
| S_W | Susceptible, previously vaccinated, protection waned (eligible for re-dosing) | Population |
| E_fast_W, E_slow_W | Latent infection, previously vaccinated, protection waned (eligible for re-dosing) | Population |
| I_PTB_SUB | Subclinical pulmonary disease (bacteriologically positive, not yet care-seeking) | Population |
| I_MIX_SUB | Subclinical concurrent pulmonary and extrapulmonary disease | Population |
| I_PTB, I_EPTB, I_MIX | Clinical active disease: pulmonary, extrapulmonary, concurrent | Population |
| T | On treatment | Population |
| R | Recovered | Population |
| CUM_PTB, CUM_EPTB, CUM_MIX | Cumulative incident cases by disease type | Cumulative counter |
| CUM_DEATHS_PTB, CUM_DEATHS_EPTB, CUM_DEATHS_MIX | Cumulative TB deaths by disease type | Cumulative counter |
| CUM_DOSES | Cumulative vaccine doses administered | Cumulative counter |
| I_PTB_SUB_VHIST, I_MIX_SUB_VHIST | Vaccinated-origin subset of the subclinical compartments | Vaccination-history counter |
| I_PTB_VHIST, I_EPTB_VHIST, I_MIX_VHIST | Vaccinated-origin subset of the clinical disease compartments | Vaccination-history counter |
| T_VHIST | Vaccinated-origin subset of the on-treatment compartment | Vaccination-history counter |
| R_VHIST | Vaccinated-origin subset of the recovered compartment; its waning is routed to S_W rather than S | Vaccination-history counter |

**Online supplemental table S2. Fixed transmission, natural-history, demographic and program parameters**

| Parameter | Value | Source / status |
|---|---|---|
| EPTB share of latent infection: base, anchor year, trend, bounds | 0·18 (2018); +1·4 pp per year; bounded to [0·05, 0·40] | India TB Report 2023/24, NTEP |
| Concurrent-disease (MIX) case fraction | 0·047 | Fixed |
| Relative infectiousness: PTB / EPTB / MIX | 1·00 / 0·30 / 0·65 | Fixed |
| Relative infectiousness of subclinical disease | 0·437260 × the disease type's own weight (calibrated, table S3) | Calibrated |
| Base detection rate: PTB / EPTB / MIX ($\tau$, /yr) | 2·5 / 1·5 / 1·8 | Fixed |
| Cure parameter / baseline cure rate (j_cure / c0) | 1·50 / 0·87 | Fixed |
| Base case fatality: PTB / EPTB / MIX ($\delta$) | 0·142 / 0·225 / 0·283 | Fixed |
| Case-fatality age multipliers, PTB (6–14 / 15–30 / 31–55 / 56–80) | 1·5 / 1·0 / 1·2 / 3·0 | Fixed |
| Case-fatality age multipliers, EPTB | 2·5 / 1·0 / 1·5 / 4·0 | Fixed |
| Case-fatality HIV multiplier (negative / positive) | 1·0 / 2·81 | Fixed |
| Case-fatality multiplier cap | 4·0 | Fixed |
| Subclinical self-cure rate / subclinical case-fatality multiplier | 0·10 /yr / 0·30 | Literature-anchored, not calibrated |
| Susceptibility multipliers: HIV (negative / positive) | 1·0 / 3·5 | Fixed |
| Susceptibility multipliers: BMI (low / normal / high) | 1·6 / 1·0 / 0·85 | Fixed |
| Relapse from recovered: PTB / EPTB / MIX (/yr) | 0·005 / 0·001 / 0·0007 | Fixed |
| Relapse from treatment: PTB / EPTB / MIX (/yr) | 0·05 / 0·03 / 0·04 | Fixed |
| Treatment other-exit rate (/yr) | 0·02 | Fixed |
| Waning of natural immunity, R → S ($\omega$, /yr) | 0·118 | Fixed |
| Background (non-TB) mortality (/yr) | 0·012 | Fixed |
| Birth rate (/yr) | 0·025 | India 2010–20 mid-range |
| Aging rates across the four age bands (/yr) | 1/9, 1/16, 1/25, 1/25 | Band widths |

| | | |
|---|---|---|
| Vaccine uptake multiplier by age (6–14 / 15–30 / 31–55 / 56–80) | 0·90 / 0·70 / 0·60 / 0·50 | Fixed |
| Vaccine uptake multiplier by SES (low / middle / high) | 0·85 / 1·00 / 1·08 | Fixed |
| Infection-blocking vaccine efficacy (ε_inf) | 0 (progression-blocking only) | Phase III trial evidence |
| CDR / TSR ceilings imposed on the logistic fits | 92% / 95% | India End TB target plus headroom |
| Vaccination program window | 2026–2050 | Fixed |
| Dose calendar | 2026, 2036, 2046 (both protection durations) | Fixed (this analysis) |
| Coverage parameter, solved so that dose 1 vaccinates exactly 700,000 of the original 1,000,000 | 0·650675 | Solved numerically |
| Age distribution (6–14 / 15–30 / 31–55 / 56–80) | 20% / 30% / 35% / 15% | 2011 India Census |
| HIV distribution (negative / positive) | 97% / 3% | NACO |
| BMI distribution (low / normal / high) | 30% / 55% / 15% | India-specific |
| SES distribution (low / middle / high) | 40% / 40% / 20% | India-specific |

**Online supplemental table S3. The 14 free parameters estimated by differential evolution, with search bounds and calibrated values**

| Parameter | Lower bound | Upper bound | Calibrated value | Note |
|---|---|---|---|---|
| Transmission coefficient (β) | 0·3 | 5 | 0·462380 | |
| PTB progression, fast pathway (K_PTB,fast, /yr) | 0·02 | 0·2 | 0·0950503 | |
| PTB progression, slow pathway (K_PTB,slow, /yr) | 0·0005 | 0·02 | 0·00429713 | |
| EPTB progression, fast pathway (K_EPTB,fast, /yr) | 0·005 | 0·15 | 0·0760187 | |
| EPTB progression, slow pathway (K_EPTB,slow, /yr) | 0·0001 | 0·02 | 0·00444768 | |
| Self-clearance, fast latent (σ_fast, /yr) | 0·01 | 0·08 | 0·0361668 | |
| Self-clearance, slow latent (σ_slow, /yr) | 0·0001 | 0·04 | 0·000986710 | |
| Fast → slow latent transition (κ, /yr) | 0·25 | 1 | 0·703945 | |
| Relative susceptibility to reinfection (ν) | 0·01 | 0·5 | 0·0111012 | Settled 0·22% above its lower bound of 0·01; the other 13 parameters are interior |
| Case-fatality multiplier (cfr_mult) | 0·5 | 3 | 0·680738 | |
| CDR trend dampening | 0·1 | 1 | 0·460717 | |
| TSR trend dampening | 0·1 | 1 | 0·584405 | |
| Subclinical → clinical progression (/yr) | 0·15 | 1·2 | 0·732830 | |
| Subclinical relative infectiousness | 0·05 | 0·6 | 0·437260 | |

**Online supplemental table S4. Bayesian evidence synthesis of vaccine efficacy: prior, likelihood and posterior on the log-hazard-ratio scale**. Vaccine effectiveness is the median, 1 − exp(log HR); the interval is the 2·5th–97·5th percentile.

| Endpoint | Stage | log HR | SE(log HR) | Vaccine effectiveness median (%) | 95% Credible Intervals [CrI] (%) |
|---|---|---|---|---|---|
| Pulmonary TB | Per-protocol prior, power-discounted (α=0·082) | -0·2169 | 0·6266 | 19·50 | -174·82 to 76·42 |
| Pulmonary TB | mITT likelihood | -0·1462 | 0·1720 | 13·60 | -21·03 to 38·32 |
| Pulmonary TB | Posterior | -0·1511 | 0·1659 | 14·02 | -19·00 to 37·88 |
| Extrapulmonary TB | Per-protocol prior, power-discounted (α=0·082) | -0·7012 | 1·2314 | 50·40 | -453·87 to 95·56 |
| Extrapulmonary TB | mITT likelihood | -0·5499 | 0·3243 | 42·30 | -8·93 to 69·44 |
| Extrapulmonary TB | Posterior | -0·5597 | 0·3136 | 42·86 | -5·63 to 69·09 |
| All TB (composite) | Per-protocol prior, power-discounted (α=0·082) | -0·2408 | 0·5782 | 21·40 | -144·03 to 74·68 |
| All TB (composite) | mITT likelihood | -0·1851 | 0·1584 | 16·90 | -13·34 to 39·07 |
| All TB (composite) | Posterior | -0·1890 | 0·1527 | 17·22 | -11·66 to 38·63 |

**Online supplemental table S5. Sensitivity to the vaccine-effectiveness clipping convention (1,000 iterations, 10-year protection, dynamic baseline, single-arm Monte Carlo routine)**.

| Convention | Mean cases averted | Median cases averted | 95% UI, cases averted | Mean DALYs averted | Iterations net-negative (%) |
|---|---|---|---|---|---|
| clipped (VE>=0; primary) | 5,758 | 5,669 | 170 to 11,169 | 10,377 | 0 |
| untruncated (sensitivity) | 5,438 | 5,669 | -2,250 to 11,169 | 9,887 | 7·2 |

**Online supplemental table S6. Vaccination schedule, eligible population and realized coverage at each dose event, per 1,000,000 population targeted**

| Protection duration | Dose event | Year | Eligible population (n) | Vaccinated (n) | Realized coverage of eligible | % of the original 1,000,000 | Cumulative doses |
|---|---|---|---|---|---|---|---|
| 3-year | Initial vaccination | 2026 | 1,075,806 | 700,000 | 65·1% | 70·0% | 700,000 |
| 3-year | First booster | 2036 | 568,344 | 369,807 | 65·1% | 37·0% | 1,069,807 |
| 3-year | Second booster | 2046 | 448,198 | 291,631 | 65·1% | 29·2% | 1,361,438 |
| 10-year | Initial vaccination | 2026 | 1,075,806 | 700,000 | 65·1% | 70·0% | 700,000 |
| 10-year | First booster | 2036 | 569,261 | 370,403 | 65·1% | 37·0% | 1,070,403 |
| 10-year | Second booster | 2046 | 451,385 | 293,705 | 65·1% | 29·4% | 1,364,108 |

**Online supplemental table S7. Cost and benefit components by dose count and protection duration, discounted, per 1,000,000 population targeted in US$**

| Protection duration | Dose schedule | Doses given (n) | Vaccination cost (95% UI) | Treatment + hospital savings (95% UI) | Productivity savings (95% UI) | Value-of-statistical-life benefit (95% UI) | Total benefit (95% UI) |
|---|---|---|---|---|---|---|---|
| 3-year | One dose (2026) | 700,000 | 1,925,000 (1,925,000–1,925,000) | 1,363,205 (63,369–2,633,206) | 3,204,345 (123,525–6,531,755) | 9,540,191 (436,405–18,549,764) | 14,107,741 (623,299–27,720,152) |
| 3-year | Two doses (2026, 2036) | 1,070,088 | 2,678,962 (2,678,410–2,679,571) | 1,891,310 (92,129–3,615,508) | 4,390,744 (179,532–8,837,619) | 13,263,965 (637,574–25,483,499) | 19,546,019 (909,235–37,744,266) |
| 3-year | Three doses (2026, 2036, 2046) | 1,362,004 | 3,119,531 (3,118,565–3,120,588) | 2,146,781 (107,461–4,080,027) | 4,952,115 (208,747–9,873,766) | 15,066,543 (747,744–28,696,536) | 22,165,440 (1,063,952–42,597,845) |
| 10-year | One dose (2026) | 700,000 | 1,925,000 (1,925,000–1,925,000) | 3,842,918 (132,696–7,090,393) | 8,856,225 (259,914–17,259,995) | 27,217,359 (927,924–50,637,335) | 39,916,502 (1,320,048–75,085,435) |
| 10-year | Two doses (2026, 2036) | 1,071,466 | 2,681,769 (2,679,671–2,683,775) | 5,230,492 (186,106–9,608,871) | 11,964,587 (363,810–23,123,678) | 37,210,982 (1,309,216–68,528,328) | 54,406,061 (1,858,449–100,890,739) |
| 10-year | Three doses (2026, 2036, 2046) | 1,366,144 | 3,126,506 (3,122,985–3,129,851) | 5,544,758 (199,284–10,179,176) | 12,654,060 (388,837–24,397,361) | 39,226,439 (1,395,187–72,191,836) | 57,425,258 (1,982,660–105,850,622) |

Benefit–cost ratio falls as doses are added — 20·7, 20·3 and 18·4 under 10-year protection for one, two and three doses — because each additional dose event costs the same per recipient but averts less. Median gross incremental cost-effectiveness ratio moves the other way, from US$286 to US$295 to US$329. Both remain far inside conventional willingness-to-pay thresholds at every dose count (figures S10).

**Online supplemental table S8. Health-economic and disability parameters**

| Parameter | Value | Source / status |
|---|---|---|
| Disability weight: PTB / EPTB / MIX | 0·333 / 0·272 / 0·5144 | PTB: Salomon 2015 (GBD 2013); EPTB: central value within a 0·225–0·333 range; MIX derived as 1 − (1 − 0·333)(1 − 0·272) |
| Disease duration: PTB / EPTB / MIX (years) | 0·5 / 0·75 / 0·75 | Fixed |
| Residual life expectancy by age band (years) | 62·0 / 50·0 / 29·0 / 11·0 | India life tables |
| Discount rate | 3% per year | Indian Reference Case |
| GDP per capita, India | US$2,694·70 | World Bank |
| Willingness-to-pay thresholds | 1× GDP US$2,695; 3× GDP US$8,084; Ochalek US$500 per DALY | WHO-CHOICE; Ochalek et al. |
| Value of a statistical life: central value and sensitivity grid | US$80,000; US$40,000 / 80,000 / 120,000 / 160,000 / 300,000 / 416,000 | LMIC literature |
| Vaccine cost per dose event: procurement + administration | US$0·75 + US$2·00 = US$2·75 | Fixed |
| Treatment cost: PTB / EPTB / MIX | US$500 / 800 / 1,000 | Fixed |
| Hospitalization cost: PTB / EPTB / MIX | US$2,000 / 3,500 / 4,500 | Fixed |
| Productivity loss: PTB / EPTB / MIX | US$3,000 / 3,500 / 4,000 | Fixed |

**Online supplemental table S9. Sensitivity to the assumed trend in the extrapulmonary share of infection**

| EPTB share trend (pp per year) | EPTB share of incidence in 2050 (%) | Total incidence in 2050 (per 100,000) | TB mortality in 2050 (per 100,000) | $R^2$, incidence | $R^2$, mortality |
|---|---|---|---|---|---|
| 0·0 | 17·6 | 124·2 | 13·93 | 0·6737 | 0·6520 |
| 0·7 | 36·7 | 105·5 | 14·09 | 0·8439 | 0·7541 |
| 1·4 | 36·8 | 100·7 | 13·51 | 0·9101 | 0·7679 |
| 2·1 | 36·8 | 99·9 | 13·42 | 0·8765 | 0·6857 |

**Online supplemental table S10. Calibration quality**

| Target | $R^2$ | RMSE (per 100,000) | MAPE | Mean bias (per 100,000) |
|---|---|---|---|---|
| Incidence, dynamic baseline | 0·9101 | 4·55 | 1·91% | -1·11 |
| Incidence, static baseline | 0·7300 | 7·88 | 3·03% | -6·42 |
| Mortality, dynamic baseline | 0·7679 | 1·16 | 3·45% | -0·06 |
| Mortality, static baseline | 0·4268 | 1·83 | 5·34% | -0·69 |
| Case-detection-rate logistic fit (Layer 1) | 0·7887 | — | — | — |
| Treatment-success-rate logistic fit (Layer 1) | 0·9027 | — | — | — |

Mean bias is model minus observed: negative values indicate under-prediction, equivalent to positive observed-minus-model residuals in figure 2C of the main document

**Online supplemental table S11. Marginal (incremental) contribution of each dose event, estimated by paired Monte Carlo (the same vaccine-effectiveness draw is used in the compared arms), dynamic baseline, per 1,000,000 population targeted**

| Protection duration | Dose event | Doses given (n) | Cases averted (n) | Deaths averted (n) | DALYs averted (n) | Net cost (US$) | BCR | Gross ICER (US$/DALY) |
|---|---|---|---|---|---|---|---|---|
| 3-year | Initial dose (2026) vs no vaccine | 700,000 | 1,106 (39–2,299) | 137 (6–270) | 2,454 (110–4,803) | 561,795 (-708,206–1,861,631) | 7·3 (0·3–14·4) | 779 (401–17,599) |
| 3-year | First booster (2036), incremental | 369,807 | 529 (24–1,039) | 70 (4–131) | 896 (47–1,680) | 225,857 (-224,487–724,651) | 7·2 (0·4–13·5) | 823 (449–16,077) |
| 3-year | Second booster (2046), incremental | 291,631 | 325 (16–618) | 44 (2–79) | 357 (18–644) | 185,098 (-19,147–427,365) | 5·9 (0·3–10·8) | 1,191 (685–24,601) |
| 10-year | Initial dose (2026) vs no vaccine | 700,000 | 3,306 (91–6,553) | 427 (14–802) | 6,751 (225–12,663) | -1,917,918 (-5,165,393–1,792,304) | 20·7 (0·7–39·0) | 286 (152–8,653) |
| 10-year | First booster (2036), incremental | 370,403 | 1,508 (48–2,842) | 205 (7–375) | 2,304 (82–4,191) | -630,805 (-1,759,583–708,693) | 19·1 (0·7–34·9) | 318 (181–1,899) |
| 10-year | Second booster (2046), incremental | 293,705 | 404 (14–742) | 50 (1–88) | 408 (10–721) | 130,471 (-110,159–435,360) | 6·8 (0·2–12·1) | 1,051 (619–42,870) |

**Online supplemental table S12. Median benefit–cost ratio across the value-of-statistical-life grid**

| Protection duration | Dose schedule | US$40,000 | US$80,000 | US$120,000 | US$160,000 | US$300,000 | US$416,000 |
|---|---|---|---|---|---|---|---|
| 3-year | One dose (2026) | 4·88 | 7·36 | 9·86 | 12·36 | 21·14 | 28·37 |
| 3-year | Two doses (2026, 2036) | 4·85 | 7·39 | 9·89 | 12·38 | 21·10 | 28·35 |
| 3-year | Three doses (2026, 2036, 2046) | 4·74 | 7·18 | 9·63 | 12·08 | 20·63 | 27·72 |
| 10-year | One dose (2026) | 13·62 | 20·65 | 27·68 | 34·70 | 59·43 | 79·91 |
| 10-year | Two doses (2026, 2036) | 13·31 | 20·31 | 27·24 | 34·23 | 58·68 | 78·97 |
| 10-year | Three doses (2026, 2036, 2046) | 12·08 | 18·40 | 24·75 | 31·10 | 53·35 | 71·76 |

Medians over 1,000 iterations; dynamic baseline; break-even is 1·0. The central case is US$80,000.

Median benefit–cost ratio remains above break-even at every tested value of a statistical life, in every dose arm and at both protection durations. The lowest median benefit–cost ratio anywhere on the grid is 4·7, for the three-dose arm under 3-year protection at a value of a statistical life of US$40,000.

**Online supplemental table S13. Probability of benefit and of cost-effectiveness (% of 1,000 iterations)**

| Protection duration | Dose schedule | P(cases averted > 0) | P(deaths averted > 0) | P(DALYs averted > 0) | P(BCR > 1) | P(cost-effective, 1×GDP) | P(cost-effective, 3×GDP) | P(cost-effective, Ochalek US$500) | Iterations with both VE draws clipped to zero (%) |
|---|---|---|---|---|---|---|---|---|---|
| 3-year | One dose (2026) | 97·9 | 97·9 | 97·9 | 95·1 | 92·8 | 96·8 | 68·3 | 2·1 |
| 3-year | Two doses (2026, 2036) | 97·9 | 97·9 | 97·9 | 95·2 | 92·9 | 96·7 | 68·7 | 2·1 |
| 3-year | Three doses (2026, 2036, 2046) | 97·9 | 97·9 | 97·9 | 95·2 | 92·7 | 96·7 | 67·1 | 2·1 |
| 10-year | One dose (2026) | 97·7 | 97·7 | 97·7 | 97·1 | 96·3 | 97·5 | 93·1 | 2·3 |
| 10-year | Two doses (2026, 2036) | 97·7 | 97·7 | 97·7 | 97·1 | 96·3 | 97·5 | 93·0 | 2·3 |
| 10-year | Three doses (2026, 2036, 2046) | 97·7 | 97·7 | 97·7 | 96·8 | 96·3 | 97·4 | 92·3 | 2·3 |

The final column is the fraction of iterations in which both vaccine-effectiveness draws were truncated to zero, so that no vaccine effect was modeled. This is the upper bound on how far the probability of a positive health effect can fall short of 100%: every iteration with at least one non-zero draw produced a positive health gain. Small differences between the cases, deaths and DALYs columns within a row are numerical noise around zero in that group and should not be interpreted.

**Online supplemental table S14. Cases and deaths averted (PTB + EPTB combined) by demographic stratum, 10-year protection, dynamic baseline, per 1,000,000 population targeted. Each row is a marginal breakdown summed over the other three stratifying dimensions**

| Dimension | Stratum | Cases: 1 dose | Cases: 2 doses | Cases: 3 doses | Deaths: 1 dose | Deaths: 2 doses | Deaths: 3 doses |
|---|---|---|---|---|---|---|---|
| AGE | 6-14 | 323 | 423 | 436 | 44·3 | 57·8 | 59·4 |
| AGE | 15-30 | 1,023 | 1,448 | 1,538 | 81·0 | 114·7 | 120·7 |
| AGE | 31-55 | 1,180 | 1,763 | 1,934 | 134·2 | 202·1 | 218·9 |
| AGE | 56-80 | 542 | 837 | 943 | 134·9 | 210·9 | 234·0 |
| HIV | HIV-negative | 2,967 | 4,328 | 4,694 | 370·4 | 551·2 | 595·9 |
| HIV | HIV-positive | 101 | 144 | 156 | 23·9 | 34·4 | 37·0 |
| BMI | Low BMI | 1,136 | 1,655 | 1,792 | 143·9 | 213·4 | 230·3 |
| BMI | Normal BMI | 1,549 | 2,258 | 2,451 | 200·5 | 297·9 | 322·1 |
| BMI | High BMI | 383 | 559 | 607 | 50·0 | 74·3 | 80·4 |
| SES | Low SES | 1,159 | 1,637 | 1,761 | 147·8 | 211·9 | 227·3 |
| SES | Middle SES | 1,307 | 1,927 | 2,095 | 168·0 | 252·5 | 273·6 |
| SES | High SES | 602 | 908 | 994 | 78·5 | 121·1 | 132·0 |

Means over 1,000 iterations. Each row is a marginal breakdown summed over the other three stratifying dimensions, not a full cross-tabulation.

Impact concentrates in working-age adults: under 10-year protection with all three doses the 31–55-year band accounts for 39·9% of combined pulmonary and extrapulmonary cases averted, the 15–30-year band 31·7%, the 56–80-year band 19·4% and the 6–14-year band 9·0%. Because vaccine effectiveness was not modeled as stratum-varying, these patterns reflect where tuberculosis burden and modeled uptake fall rather than any differential biological effect of the vaccine. The low-socioeconomic-status stratum shows fewer absolute cases averted than the middle stratum despite carrying higher disease burden, because modeled uptake there is lower (0·85 against 1·00). Figure S11 shows the breakdown for cases and for deaths.

**Online supplemental table S15. Reproducibility record for the run that produced every number reported here**

| Item | Value |
|---|---|
| Model version | v25.4_3doses_booster_dose (tb_model_v25_3doses_booster_dose_WHO_FIT_CHECKLIST_FULLCAL.py) |
| Model file SHA-256 | 9ebc1a2b7d6fe4ac03d02b41c6cfe0fb1ce380bb6d95d59369073ca99be97464 |
| Calibration cache | calibration_cache_v25_4_corrected_relapse.pkl |
| Calibration cache SHA-256 | 6eb6f162aea8a2e7e116b9b2292a29ffa8cee77aaefeb015a1213879da5e1754 |
| Reference workbook SHA-256 | 740a0ee4f7f40a1c270eea0c2a633fb354467eb9287eb3279cdd36448a78e4eb |
| Output directory | tb_model_v25_3doses_Sep19_main_1000_outputs |
| Monte Carlo iterations per scenario | 1,000 |
| Vaccine-effectiveness clipping convention | Clipped at zero (primary) |
| Clipping-sensitivity iterations | 1,000 |
| Dose calendar | 2026, 2036, 2046 (both protection durations) |
| Coverage parameter (solved) | 0·650675 |
| Target vaccinated at dose 1 | 700,000 |
| State space | 30 states × 72 strata = 2,160 compartments |
| Differential-evolution convergence criterion met | No |
| Differential-evolution best loss | 0·258424 |
| Differential-evolution wall time | 10·07 h (200 generations; iteration limit reached) |
| Parameters at a search bound | One — relative susceptibility to reinfection (ν), 0·22% above its lower bound of 0·01; the other 13 are interior |
| Seeding | numpy SeedSequence combining a base seed, a per-scenario hash and the iteration index; the base seed is matched across dose-count arms within each protection duration so that incremental effects are paired |
| Random-effects propagated in the base case | Vaccine-effectiveness posterior only; calibration-parameter uncertainty is not jointly propagated |

*Every number reported in the main text, tables and figures comes from this single run.*

1. Prem K, Cook AR, Jit M. Projecting social contact matrices in 152 countries using contact surveys and demographic data. *PLoS Comput Biol* 2017;13(9):e1005697. doi: 10.1371/journal.pcbi.1005697 [published Online First: 20170912]

### Online Supplementary Figures for:

### A disaggregated dynamic model of pulmonary and extrapulmonary tuberculosis for vaccine and booster-schedule evaluation: VPM1002 in India as a case study

M Revathy, Vidya Niranjan, Akshay Uttarkar, Varun Swaminathan, Alessandro Carrese, and Aditi Bhargava

**Online supplemental figure S1. Four-parameter logistic (curve-bending) fits to India's case-detection and treatment-success rates, 2015–2024.**

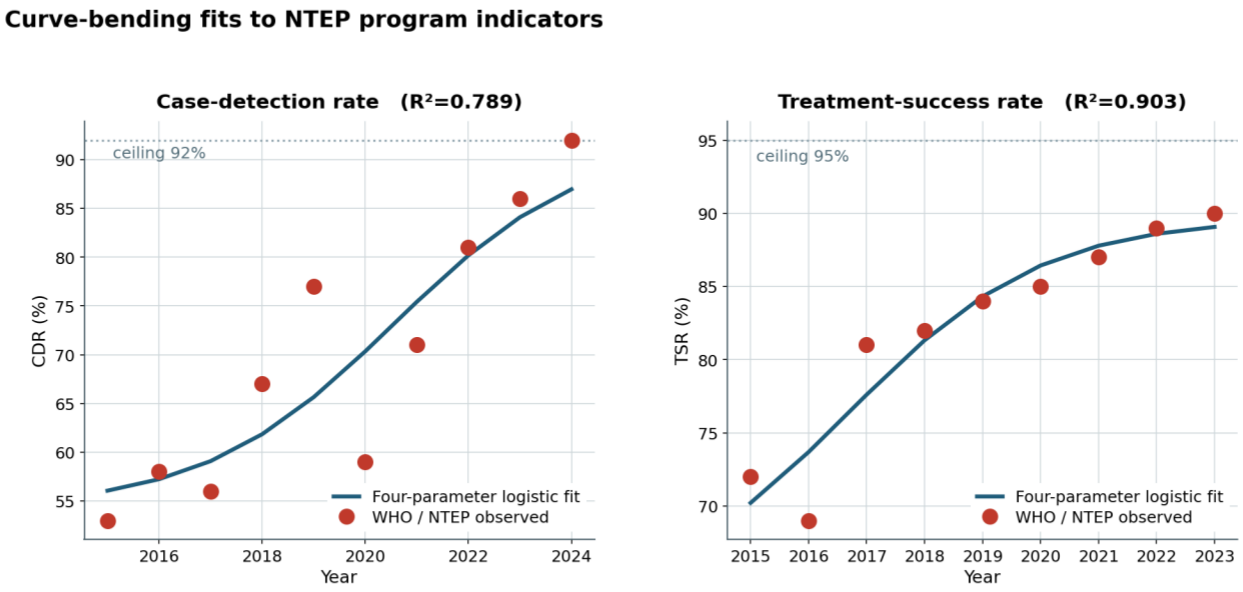


Observed points are WHO and NTEP values; fitted curves are four-parameter logistic functions with the calibration origin at 2015. Upper asymptotes are hard ceilings (92% for case detection, 95% for treatment success) rather than freely estimated values. Case detection rises from a fitted lower asymptote of 54·3% toward the ceiling at 0·54 per year ($R^2$=0·789); the fit is materially looser over 2015–2020, where the observed series swings from 53% to 77% including a 2020 dip consistent with COVID-19 service disruption while the fitted curve sits near its lower asymptote, and both series tighten from 2021 onward. Treatment success rises from a fitted 70·2% in 2015 to 89·1% in 2023 ($R^2$=0·903), the last year for which that indicator was available.

**Online supplemental figure S2. Bayesian evidence synthesis of the trial's vaccine efficacy.**

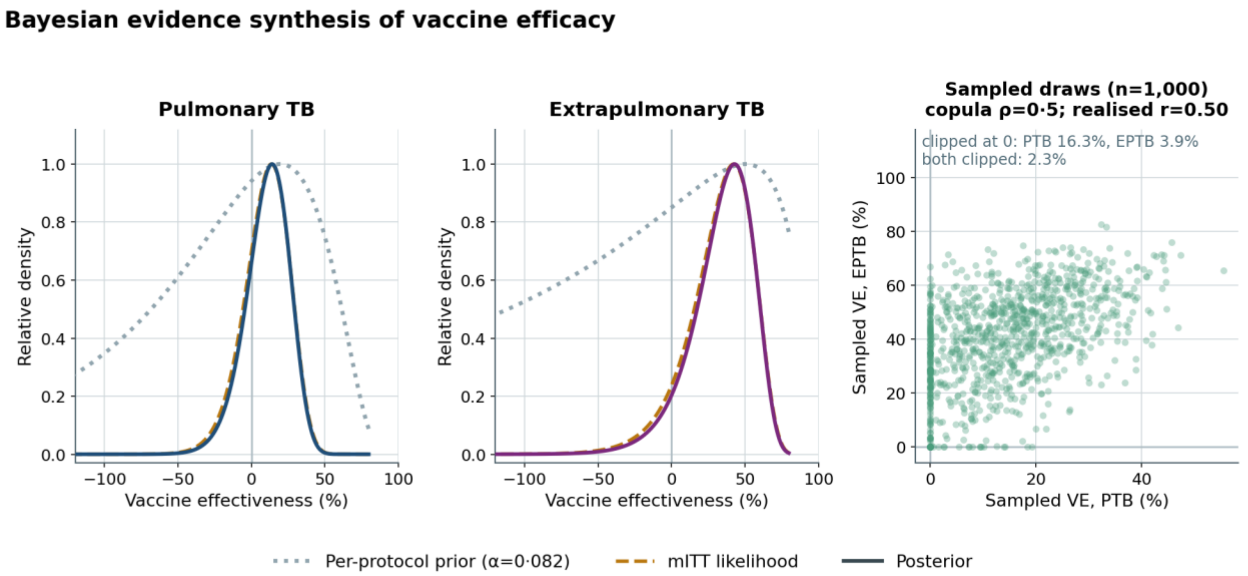


Left and center panels: prior (per-protocol, power-discounted at α=0·082), likelihood (modified intention-to-treat) and posterior densities for pulmonary and extrapulmonary disease, plotted on the vaccine-effectiveness scale and normalized to a common peak height. Because the prior is heavily discounted for its approximately 92% participant overlap with the likelihood, the posterior sits almost on top of the likelihood; this is the intended behavior of a power prior at this weight, not a failure of the prior to inform the update. Right panel: scatter plot showing 1,000 sampled draws used in every reported scenario, representing the Gaussian copula correlation (nominal ρ=0·5, realized r=0·50) and the truncation of negative draws at zero, which produces the visible accumulation along both axes. Posterior median effectiveness is 14·0% for pulmonary and 42·9% for extrapulmonary disease (table S4).

**Online supplemental figure S3. Cost and benefit structure by number of doses given.**

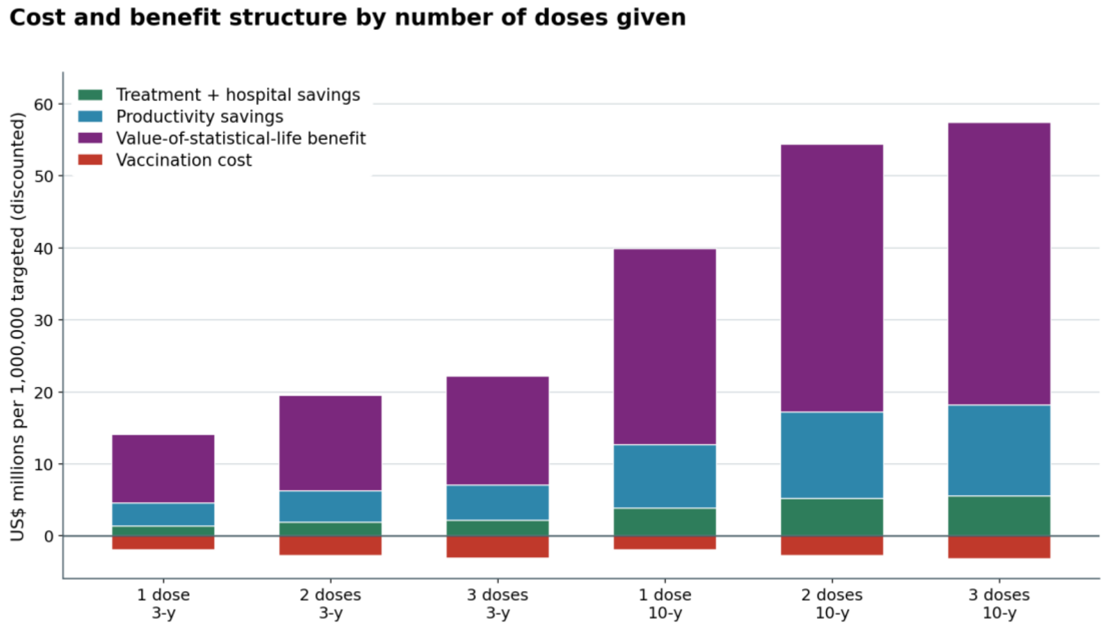


Stack plots showing discounted US$ millions per 1,000,000 population targeted with dynamic baseline and means over 1,000 iterations. Vaccination cost appears negative because administration costs US$2·00 per dose event against US$0·75 for procurement, and because a three-dose schedule delivers approximately 1·36 million doses per 1,000,000 people targeted, administration rather than procurement dominates program cost.

**Online supplemental figure S4. Sensitivity to the assumed trend in the extrapulmonary share of infection.**

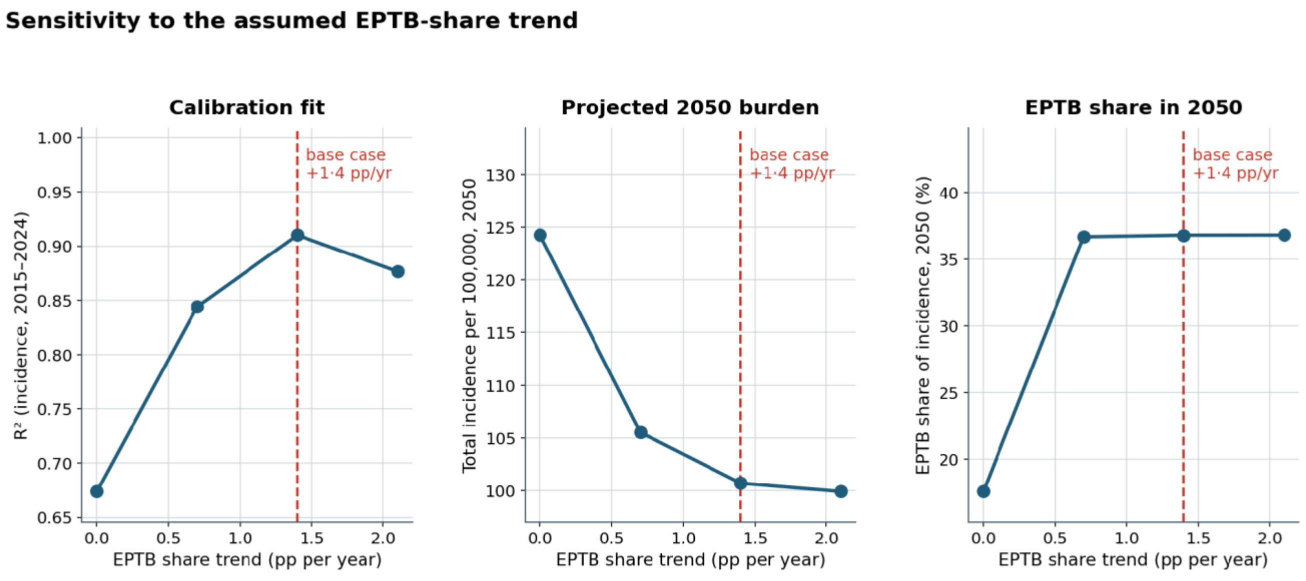


Line plots showing four candidate trends in the extrapulmonary share of latent infection tested (0·0, 0·7, 1·4 and 2·1 percentage points per year). The base-case trend of +1·4 maximizes the incidence fit among those tested. Setting the trend to zero degrades the incidence $R^2$ from 0·910 to 0·674 and the mortality $R^2$ from 0·768 to 0·652, and gives a 2050 extrapulmonary share of 17·6% against 36·8% in the base case. A rising extrapulmonary share is therefore required to reproduce the observed series rather than imposed on it, although the specific trend value is selected rather than jointly estimated and the projected extrapulmonary trajectory depends on it (table S9).

**Online supplemental figure S5. Effect of the vaccine-effectiveness truncation convention.**

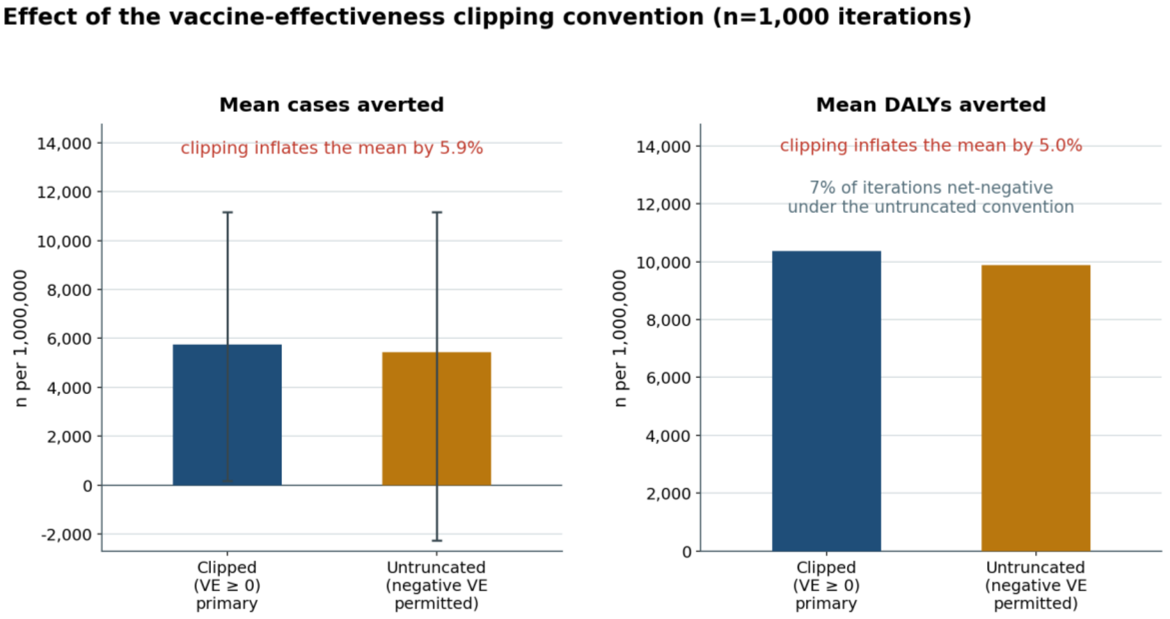


Bar graphs show 1,000 iterations, 10-year protection, dynamic baseline, single-arm Monte Carlo routine. Truncating negative effectiveness draws at zero, the base-case convention, inflates mean cases averted by 5·9% and mean DALYs averted by 5·0% relative to the untruncated posterior, under which 7·2% of iterations are net-negative and the lower bound of the uncertainty interval on cases averted falls below zero. Medians are identical under the two conventions, because truncation affects only the lower tail. Both rows draw on the same sampled effectiveness values and differ only in whether each is truncated, so the comparison is paired within iterations; only the ratio between the two conventions is interpreted, not the absolute level (table S5).

**Online supplemental figure S6. Monte Carlo output distributions for the full three-dose program, 10-year protection, dynamic baseline.**

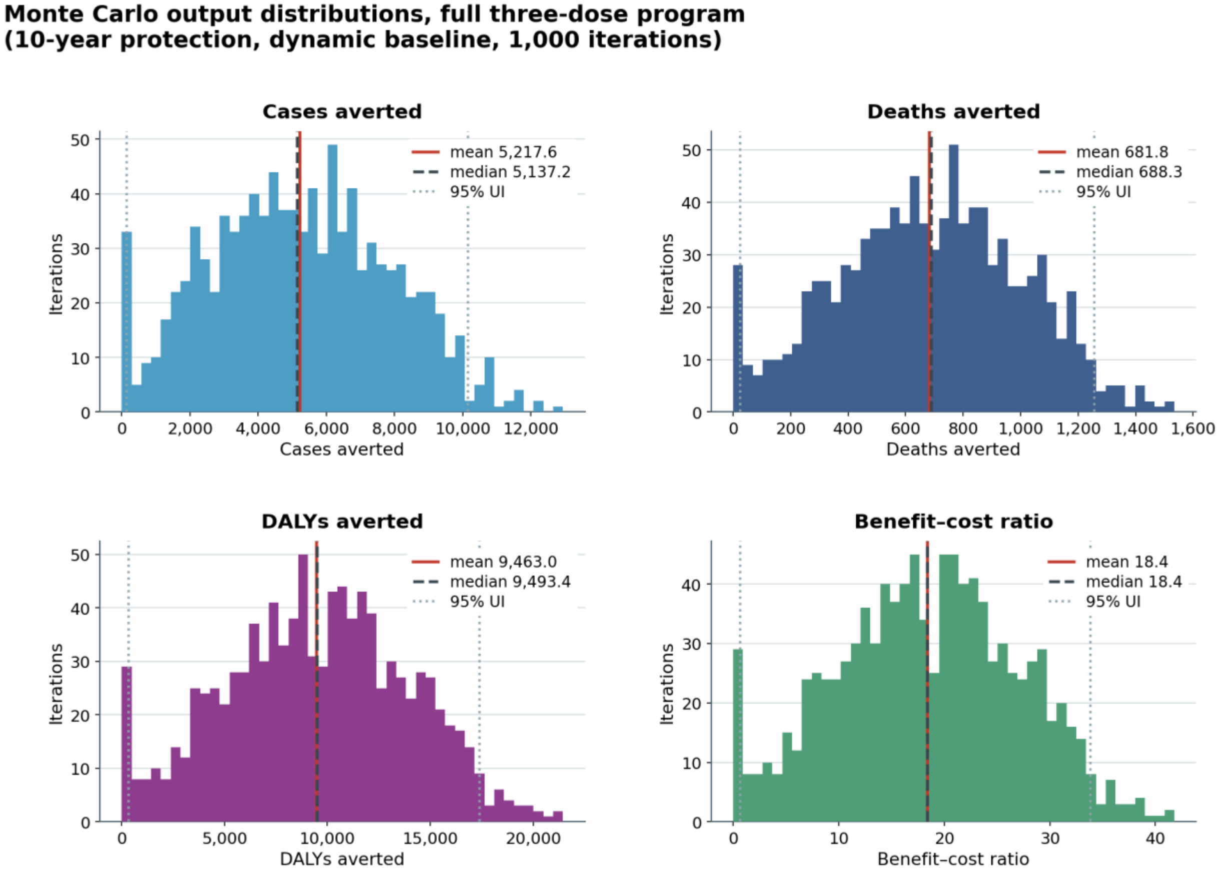


Histograms showing dose-schedule run for 1,000 iterations. All four distributions are right skewed. The spike at zero in the health-outcome panels comprises the 2·3% of iterations in which both vaccine-effectiveness draws were truncated to zero.

**Online supplemental figure S7. Disability-adjusted life-years averted per 1,000,000 population targeted.**

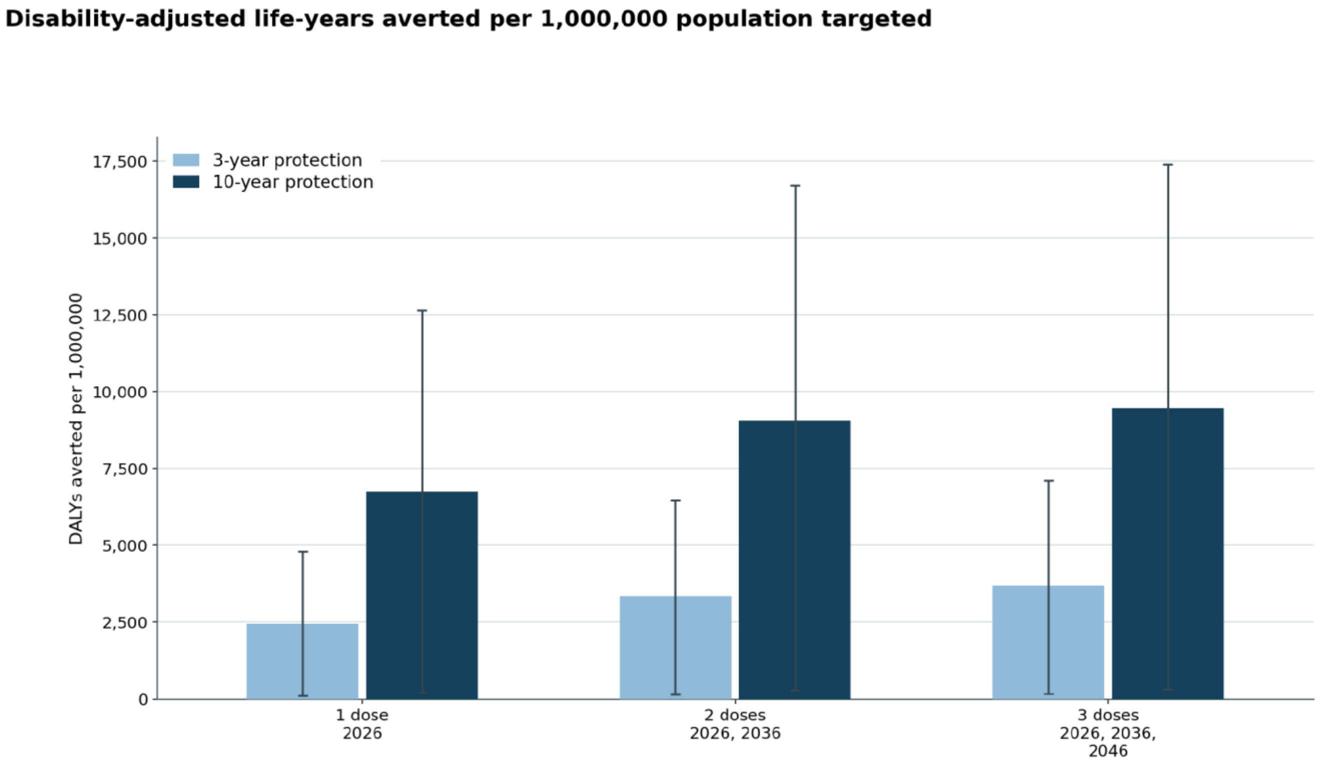


Bar graphs showing the number of doses given and protection duration, on the fixed 2026/2036/2046 dose calendar (dose-schedule run, dynamic baseline). Means with 95% uncertainty intervals over 1,000 iterations; discounted at 3% per year. Values are cumulative: the three-dose bar already contains the effect of the first and second doses. Ten-year protection increases DALYs averted approximately at 2·6–2·8-fold at every dose count, at an almost identical number of doses delivered.

**Online supplemental figure S8. Disease-type composition of averted burden, by number of doses given and protection duration.**

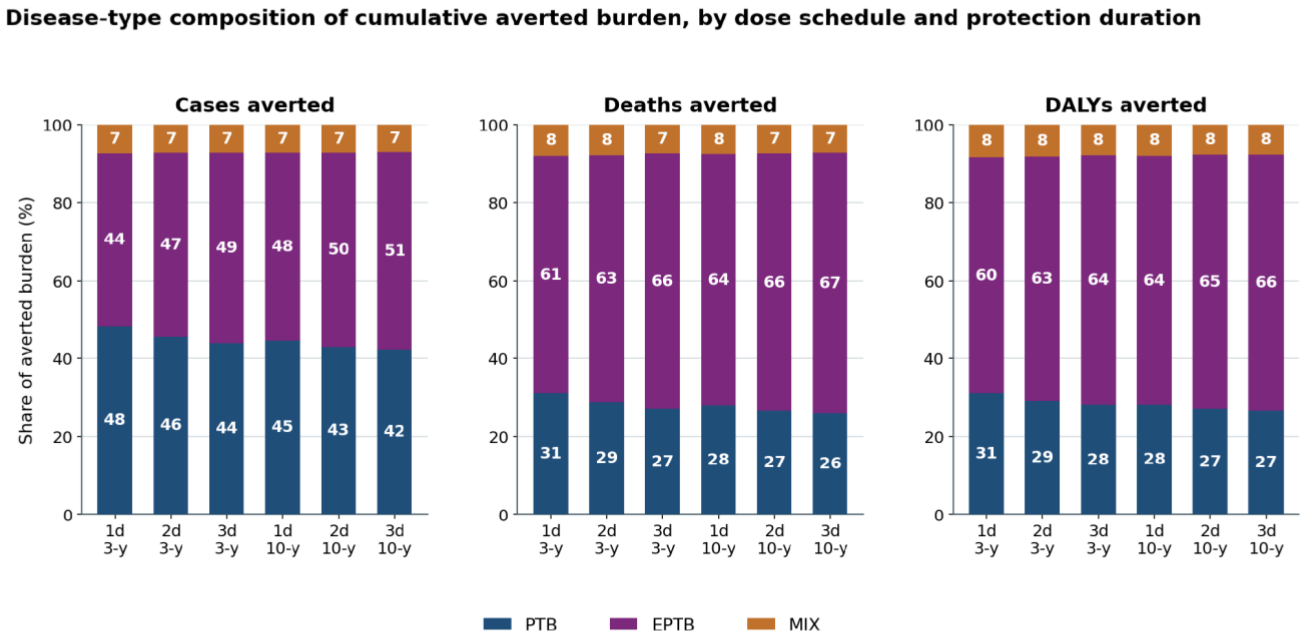


Stacked shares of the mean total across the three disease types; dynamic baseline; 1,000 iterations. Labels are percentages. The extrapulmonary share of averted deaths and DALYs rises with each additional dose and with longer protection, and exceeds the pulmonary share throughout; for averted cases the extrapulmonary share also exceeds the pulmonary share in every arm except the single 3-year dose.

**Online supplemental figure S9. Marginal contribution of each dose event, computed as a paired difference on identical vaccine-effectiveness draws.**

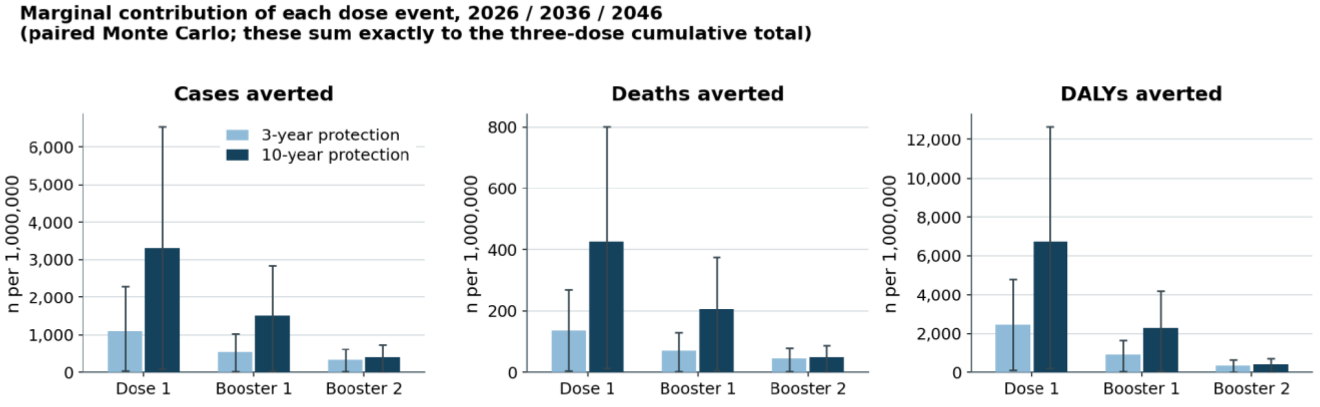


Column charts showing dynamic baseline; means with 95% uncertainty intervals over 1,000 iterations. Within each protection duration the one-, two- and three-dose arms share the same random seed, so each bar is a true marginal effect rather than a difference of two independently noisy summary statistics. Under 10-year protection the first booster contributes 1,508 averted cases and the second only 404, in part because six of the second booster's ten protected years fall beyond the 2050 horizon.

**Online supplemental figure S10. Benefit–cost ratio by number of doses given.**

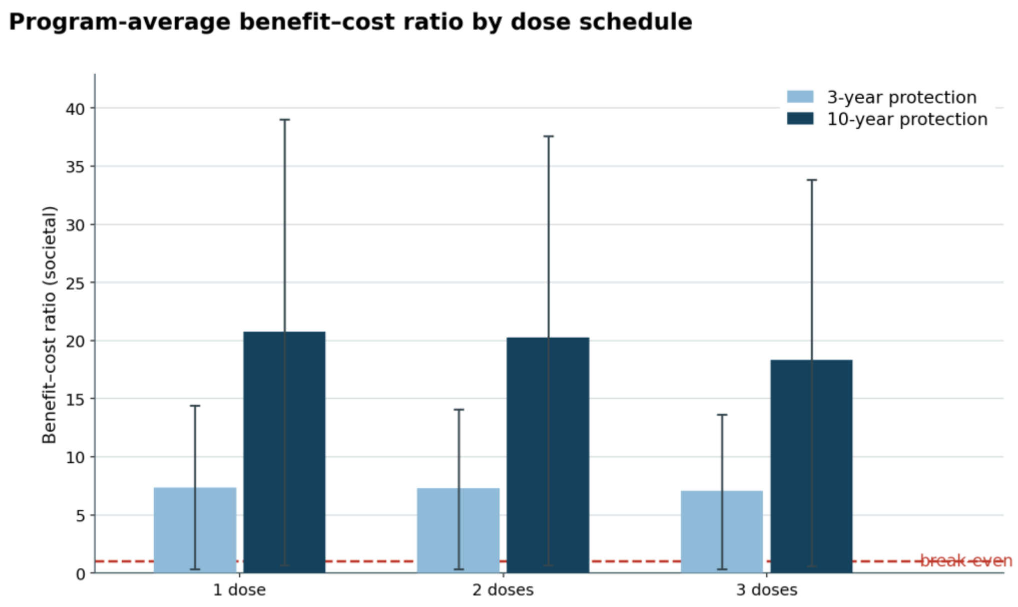


The ratio shown in the column charts is the program average for each complete schedule, not the marginal ratio for the last dose added; marginal ratios are in table S11. Societal perspective, value of a statistical life US$80,000; dynamic baseline; means with 95% uncertainty intervals over 1,000 iterations; break-even marked at 1·0. The ratio falls with each additional dose: 20·7, 20·3 and 18·4 under 10-year protection, because each booster reaches a smaller eligible pool than the dose before it while costing the same per dose delivered. The lower bound of the uncertainty interval falls below break-even in every arm.

**Online supplemental figure S11. Cases and deaths averted by demographic stratum and number of doses given, 10-year protection.**

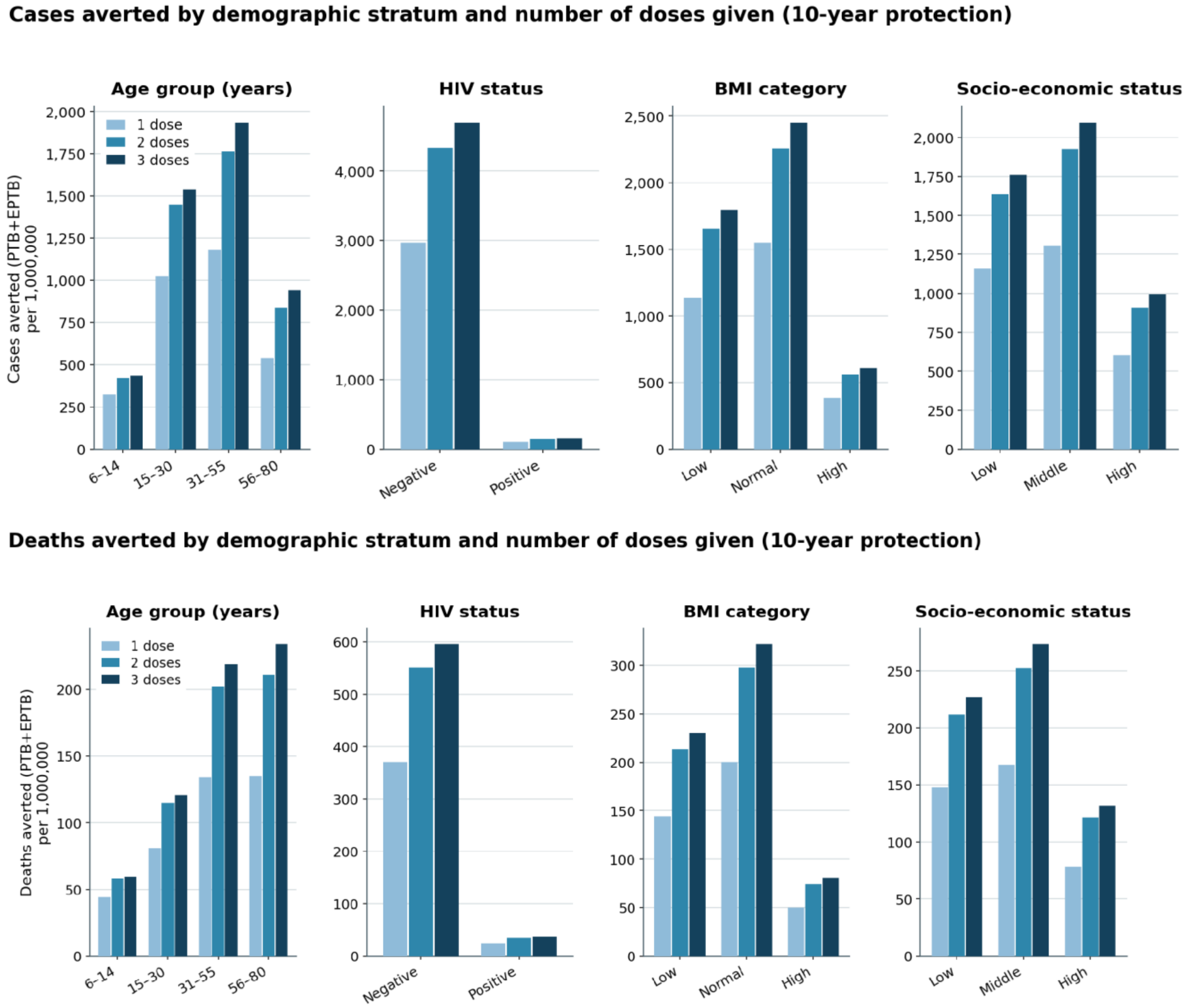


Upper panel: Column charts depicting combined pulmonary and extrapulmonary cases averted per 1,000,000 population targeted; dynamic baseline; means over 1,000 iterations. Each panel is a marginal breakdown by one stratifying dimension, summed over the other three, not a full cross-tabulation. Because vaccine effectiveness was not modeled as stratum-varying, these patterns reflect where tuberculosis burden and modeled uptake fall, not any differential biological effect of the vaccine. The low-socioeconomic-status stratum shows fewer absolute cases averted than the middle stratum despite carrying higher disease burden, because modeled uptake there is lower (0·85 against 1·00). Lower panel: Combined pulmonary and extrapulmonary deaths averted per 1,000,000 population targeted; dynamic baseline; means over 1,000 iterations.
